# Analyses of vaccine-specific circulating and bone marrow-resident B cell populations reveal benefit of delayed vaccine booster dosing with blood-stage malaria antigens

**DOI:** 10.1101/2023.03.17.23287040

**Authors:** JR Barrett, SE Silk, CG Mkindi, KM Kwiatkowska, MM Hou, AM Lias, WF Kalinga, IM Mtaka, K McHugh, M Bardelli, H Davies, LDW King, NJ Edwards, VS Chauhan, P Mukherji, S Rwezaula, CE Chitnis, AI Olotu, AM Minassian, SJ Draper, CM Nielsen

## Abstract

We have previously reported primary endpoints of a clinical trial testing two vaccine platforms for delivery of *Plasmodium vivax* malaria DBPRII: viral vectors (ChAd63, MVA) and protein/adjuvant (PvDBPII with 50µg Matrix-M™ adjuvant). Delayed boosting was necessitated due to trial halts during the pandemic and provides an opportunity to investigate the impact of dosing regimens. Here, using flow cytometry – including agnostic definition of B cell populations with the clustering tool CITRUS – we report enhanced induction of DBPRII-specific plasma cell and memory B cell responses in protein/adjuvant versus viral vector vaccinees. Within protein/adjuvant groups, delayed boosting further improved B cell immunogenicity as compared to a monthly boosting regimen. Consistent with this, delayed boosting also drove more durable anti-DBPRII serum IgG. In an independent vaccine clinical trial with the *P. falciparum* malaria RH5.1 protein/adjuvant (50µg Matrix-M™) vaccine candidate, we similarly observed enhanced circulating B cell responses in vaccinees receiving a delayed final booster. Notably, a higher frequency of vaccine-specific (putatively long-lived) plasma cells were detected in the bone marrow of these delayed boosting vaccinees by ELISPOT and correlated strongly with serum IgG.

Finally, following controlled human malaria infection with *P. vivax* parasites in the DBPRII trial, *in vivo* growth inhibition was observed to correlate with DBPRII-specific B cell and serum IgG responses. In contrast, the CD4+ and CD8+ T cell responses were impacted by vaccine platform but not dosing regimen, and did not correlate with *in vivo* growth inhibition in a challenge model. Taken together, our DBPRII and RH5 data suggest an opportunity for dosing regimen optimisation in the context of rational vaccine development against pathogens where protection is antibody-mediated.

## Introduction

The roll-out of various SARS-CoV-2 vaccines during the COVID-19 pandemic highlighted the importance of understanding the immunological significance of booster dosing timing in order to maximise protective efficacy. Increased peak serum antibody concentrations were observed with delayed booster regimens in both viral vector [1; 2] and mRNA [3] delivery platforms, indicating potential opportunities to maximise humoral immunity through optimisation of antigen delivery timing. The impact of delayed booster regimens has also been explored in more depth by earlier work from the *Plasmodium falciparum* malaria field with both the blood-stage malaria vaccine candidate RH5 [4; 5] and the pre-erythrocytic vaccine candidate RTS,S (now WHO-approved as Mosquirix; [6; 7; 8; 9; 10]). Notably, the impact of delayed booster dosing on vaccine-specific serum IgG durability has been published less widely; the RH5 trials remain the only example of improvements in durability through modifications in booster dosing regimens [4; 5]. We have previously speculated on the underlying biological mechanisms and reasons for discrepancies between the RTS,S and RH5 trials [5], and these questions clearly require further investigation. Furthermore, it is critical to confirm whether the delayed booster phenomenon is a broader immunological principle, or an anomaly restricted to the RH5.1/AS01 vaccine candidate.

Here, we present analyses of B cell, T cell and serum antibody responses to a blood-stage malaria antigen from a different species of malaria – DBPRII, *Plasmodium vivax* – and investigate the impact of delayed boosters in the context of heterologous viral vector or protein/adjuvant vaccine platforms (**Table 1**). This work builds on the initial clinical trial publication, which focused on the efficacy results of the controlled human malaria infection (CHMI) [11]. Supporting these findings, we additionally present peripheral B cell analyses (using the same B cell flow cytometry panel) from a *P. falciparum* RH5.1/Matrix-M™ vaccine trial where PBMC samples were available for exploratory analyses. For the first time in the context of malaria vaccinology, we also report on vaccine-specific bone marrow plasma cells.

**Table 1.** Vaccination regimens in DBPRII and RH5 clinical trials. Post-vaccination samples analysed in this study are following the final vaccination (FV), indicated by the highlighted cell. See Methods for further details of clinical trial design.

| Target | Platform | Dosing regimen | Timing of vaccinations<br>(m = month) |  |  |  | n<sup>s</sup> |
| --- | --- | --- | --- | --- | --- | --- | --- |
|  |  |  | Dose 1 | Dose 2 | Dose 3 | Dose 4 |  |
| *P. vivax*<br>DBPRII | ChAd63-DBPRII and MVA-DBPRII viral vectors | Monthly [VV-M] | 0m | 2m | - |  | 6 |
|  |  | Delayed [VV-D] | 0m | 17m | 19m |  | 2 |
|  | PvDBPII protein/Matrix-M™ | Monthly [PA-M] | 0m | 1m | 2m |  | 4 |
|  |  | Delayed [PA-D] | 0m | 1m | 14m |  | 8 |
|  |  | Delayed + booster [PA-DB] | 0m | 1m | 14m | 19m | 5 |
| *P. falciparum*<br>RH5 | RH5.1 protein/Matrix-M™ | Monthly [M] | 0m | 1m | 2m |  | 5 |
|  |  | Delayed [D] | 0m | 1m | 6m* |  | 6 |

<sup>s</sup> Reflects sample size of groups completing full vaccination regimen in clinical trials [11]. All trial participants were healthy adults. Sample sizes for individual assays are specified in figure legends.
\* Final dose of antigen (but not adjuvant) in the delayed arm of *P. falciparum* RH5 trial was also fractionated (see Methods).

Taken together, our data support delayed booster vaccination in the next phase of *P. vivax* DBPRII and *P. falciparum* RH5 vaccine development and, more broadly, support consideration of alternative dosing regimens for vaccines against a range of pathogens where protection is antibody-mediated.

## Results

### Delayed booster dosing increases induction of circulating DBPRII-and RH5-specific plasma cells and memory B cells

Using fluorophore-conjugated DBPRII protein probes to detect DBPRII-specific cells, we evaluated the capacity of the viral vector and protein/adjuvant platforms to drive a range of B cell responses (gating strategy shown in **Supplemental Figure 1A**). Within the plasma cell population (CD19+CD27+CD38+; **Figure 1A**) – which peaked 7-days after final vaccination – significantly higher frequencies of DBPRII-specific cells were observed in protein/adjuvant as compared to heterologous viral vector vaccinees at FV+7 and FV+14. Within the memory IgG+ B cell population (CD19+CD27+IgG+ [excluding plasma cells]; **Figure 1B**), frequencies of DBPRII-specific cells were significantly higher in protein/adjuvant vaccinees at all post-vaccination time points. Consistent with expected differences in kinetics of short-lived plasma cell (SLPC) and memory B cell responses, DBPRII-specific plasma cells peaked earlier at FV+7 and then declined by FV+28, whilst frequencies of DBPRII-specific memory IgG+ B cells were better maintained in peripheral blood between FV+7 and FV+28 (with a trend towards a peak at FV+14 [5]).

**Figure 1.**
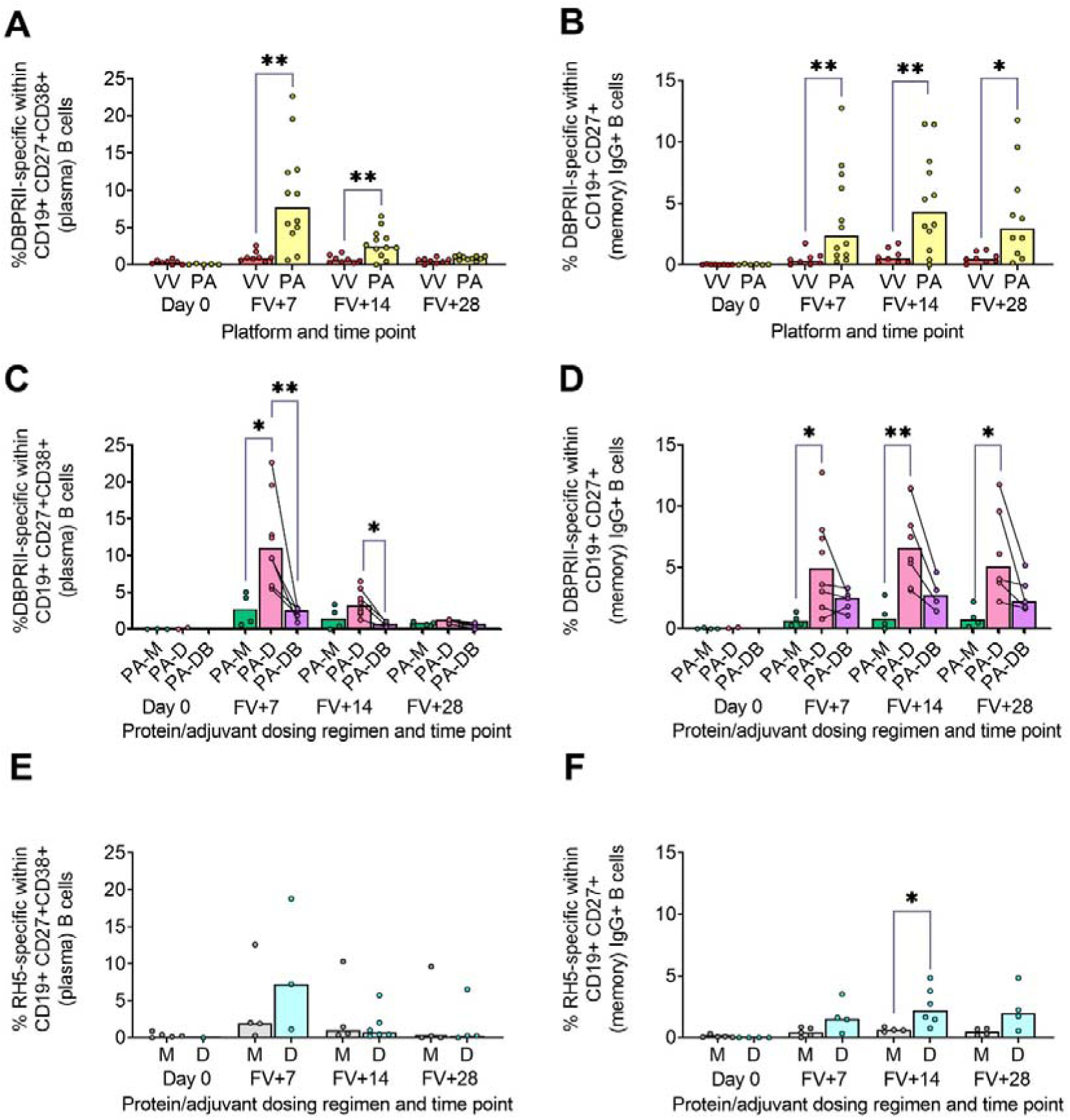
Vaccine-specific plasma cell and memory IgG+ B cell responses. PBMC from pre-vaccination (Day 0) and post-final vaccination (FV) time points were analysed for B cell responses by flow cytometry; gating strategies are as described in Methods and **Supplemental Figures 1 and 3**. Frequencies of DBPRII-specific B cells – identified by probe staining – were compared between vaccine platforms (**A-B**) or protein/adjuvant dosing regimens (**C-D**) within both plasma cell (**A, C**) or memory IgG+ B cell (**B, D**) populations. Similarly, frequencies of RH5-specific B cells were compared between protein/adjuvant dosing regimens within plasma cell (**E**) and memory IgG+ B cells (**F**). IgM+, IgA+, activated and resting memory B cell responses are shown in **Supplemental Figures 2 and 4**. VV = ChAd63-MVA viral vectors; PA = PvDBPII protein/adjuvant [PA-M and PA-D]; PA-M = PvDBPII protein/adjuvant monthly dosing; PA-D = PvDBPII protein/adjuvant delayed booster dosing; PA-DB = PvDBPII protein/adjuvant delayed booster dosing with extra booster; M = RH5.1/adjuvant monthly dosing; D = RH5.1/adjuvant delayed booster dosing. Post-vaccination comparisons were performed between DBPRII platforms (**A-B**) or RH5 dosing regimens (**E-F**) with Mann-Whitney U tests, or between PvDBPII protein/adjuvant dosing regimens by Kruskal Wallis test with Dunn’s correction for multiple comparisons (**C-D**). Sample sizes for all assays were based on sample availability; each circle represents a single sample. (**A-B**) VV/PA: Day 0 = 8/5-6, FV+7 = 8/12, FV+14 = 8/12, FV+28 = 8/10. (**C-D**) PA-M/PA-D/PA-DB: Day 0 = 3-4/2/na, FV+7 = 4/8/5, FV+14 = 4/8/4, FV+28 = 4/6/5. (**E-F**) M/D: Day 0 = 5/1-4, FV+7 = 4-5/3-4, FV+14 = 4/6, FV+28 = 4/4. PA-D vaccinees returning in the PA-DB group are connected by lines. Bars represent medians. * *p* < 0.05, ** *p* < 0.01.

We next compared DBPRII-responses between the dosing regimens of the protein/adjuvant platform. Here, we observed more robust plasma cell (**Figure 1C**) and memory IgG+ B cell (**Figure 1D**) responses following delayed booster dosing (PA-D) as compared to monthly booster dosing (PA-M). Proliferation – as indicated by intracellular Ki67 staining – was higher in plasma cells than memory B cells across all groups and time points (**Supplemental Figure 1B–C**). Differences between platforms and dosing regimens remained when we stratified between activated (CD21-CD27+) and resting (CD21+CD27+) memory IgG+ B cell populations (**Supplemental Figure 2A–D**). Interestingly, Ki67 expression was significantly higher in activated memory IgG+ B cells in PA-D at FV+7 as compared to PA-M or PA-DB, and at FV+14 as compared to PA-M (**Supplemental Figure 1C–D**). Very low memory IgA+ B cell responses were detectable (higher in protein/adjuvant than viral vector vaccinees), while responses within the IgM+ memory compartment were negligible (**Supplemental Figure 2E–H**).

We sought to validate these delayed booster dosing-mediated differences with samples from an independent clinical trial with a different cohort (Tanzanian adults) and antigen (RH5). Here, we observed comparable plasma cell and memory IgG+ B cell kinetics and differences in proliferation (**Figure 1E–F**; **Supplemental Figure 3**). Frequencies of RH5-specific cells within both populations were again higher in delayed booster dosing vaccinees but only reached statistical significance (by Mann Whitney test) for RH5-specific memory B cells at FV+14, likely related to greater intra-group variation as compared to the DBPRII vaccinees. Trends were comparable, but not significant, when activated and resting memory IgG+ B cells were analysed separately (**Supplemental Figure 4**). To note, while background with the RH5 probes was more variable than observed with the DBPRII probes here, or in previous studies with the RH5 probe protocol [5; 12], standardisation of RH5 probe gating between all samples and parent populations retains confidence in the data interpretation. No significant post-vaccination responses were observed for RH5-specific memory IgA+ or IgM+ populations (**Supplemental Figure 4**).

### Agnostically-defined B cell subsets reveal further differences in DBPRII-and RH5-specific B cell responses between vaccine platforms and dosing regimens

To supplement the more traditional approach above, clustering was next performed with CITRUS to agnostically define B cell populations for further analysis within both the DBPRII and RH5 datasets. For the DBPRII trial samples, 33 clusters were identified in CITRUS within live single (B cell-enriched) lymphocytes. Several sets of clusters had very similar marker expression patterns, resulting in consolidation for FlowJo gating strategies (see Methods). This gave a total of 7 agnostically-defined populations to reanalyse for DBPRII-specific responses (**Table 2**). Likewise, 36 clusters were identified in the RH5 trial samples, consolidating to 10 new populations for reanalysis of RH5-specific responses (**Table 2**). Populations where significant differences in antigen-specific responses were detected post-vaccination or between dosing regimens are shown in **Figure 2** (gating shown in **Supplemental Figure 5**). DBPRII “Population 2” and RH5 “Population 8” were equivalent, while all other populations identified were unique to the separate trials.

**Figure 2.**
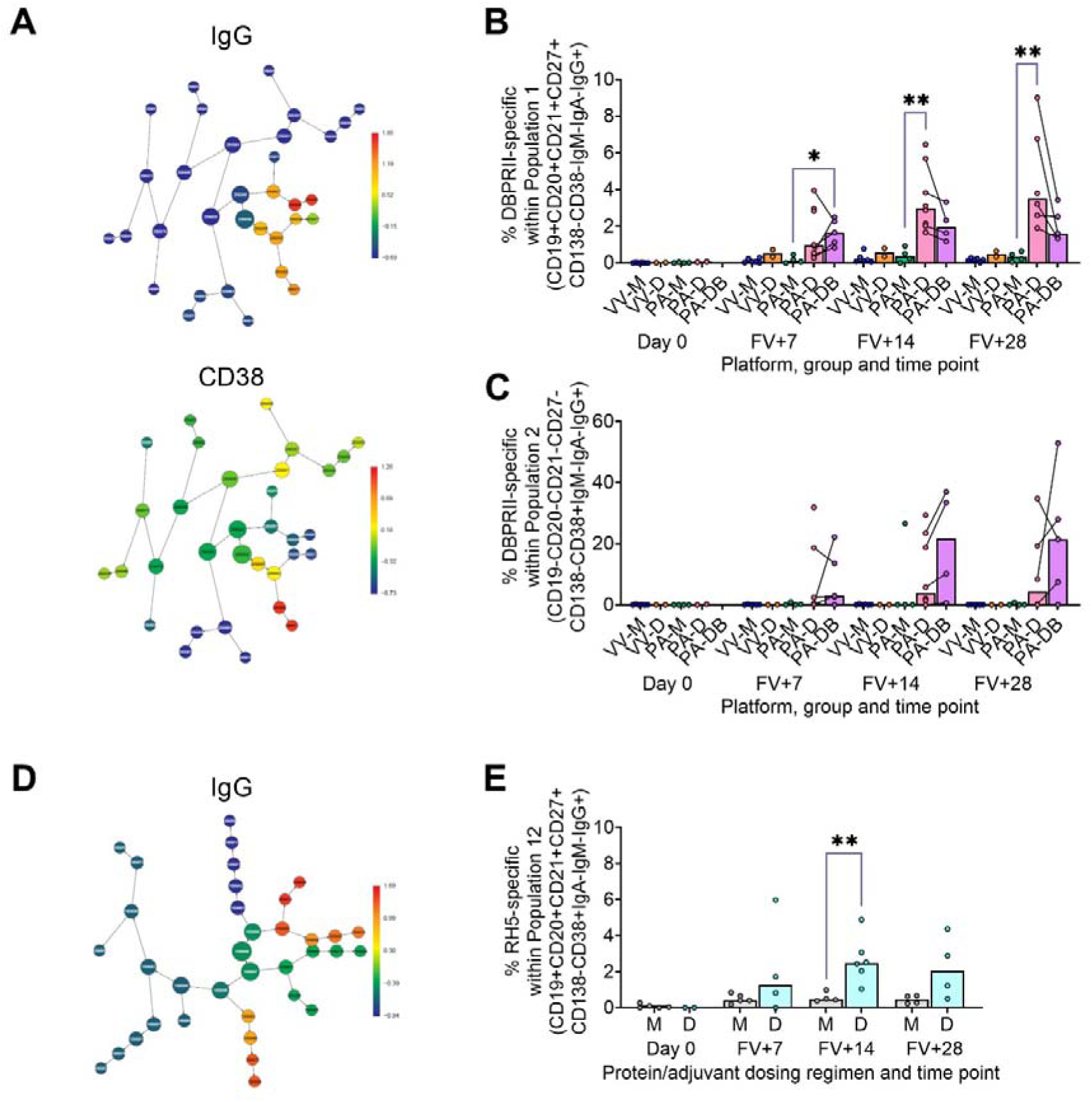
Vaccine-specific responses within agnostically-defined B cell populations using CITRUS. CITRUS was run on single live (B cell-enriched) lymphocyte flow cytometry fcs files to agnostically define the main B cell populations within either DBPRII (**A-C**) or RH5 (**D-E**) trial samples. Clusters identified by CITRUS are visualised in dendrograms (**A**, **D**), colour-coded for example markers of interest (**A**-IgG, CD38; **D**-IgG). Each node represents a cluster. Median marker expression within each cluster was used to define gating strategies for B cell populations in FlowJo, which were re-analysed for DBPRII-(**B-C**) or RH5-specific (**E**) responses through probe staining (gating shown in **Supplemental Figure 5**). See **Table 2** and **Supplemental Figures 6–7** for a full list of populations identified via CITRUS clusters for further analysis. VV-M = ChAd63-MVA viral vector monthly dosing; VV-D ChAd63-MVA delayed booster dosing; PA-M = PvDBPII protein/adjuvant monthly dosing; PA-D = PvDBPII protein/adjuvant delayed booster dosing; PA-DB = PvDBPII protein/adjuvant delayed booster dosing with extra booster; M = RH5.1/adjuvant monthly dosing; D = RH5.1/adjuvant delayed booster dosing. FV = final vaccination. Post-vaccination comparisons were performed between PvDBPII protein/adjuvant dosing regimens by Kruskal Wallis test with Dunn’s correction for multiple comparisons (**B-C**) or RH5 dosing regimens (**E**) with Mann-Whitney U tests. Sample sizes for all assays were based on sample availability; each circle represents a single sample. (**B-C**) VV-M/VV-D/PA-M/PA-D/PA-DB: Day 0 = 6/2/4/2/na, FV+7 = 6/2/4/8/5, FV+14 = 6/2/4/8/4, FV+28 = 6/2/4/6/5. (**E**) M/D: Day 0 = 5/2, FV+7 = 5/4, FV+14 = 4/6, FV+28 = 4/4. PA-D vaccinees returning in the PA-DB group are connected by lines. Bars represent medians. * *p* < 0.05, ** *p* < 0.01.

**Table 2.**
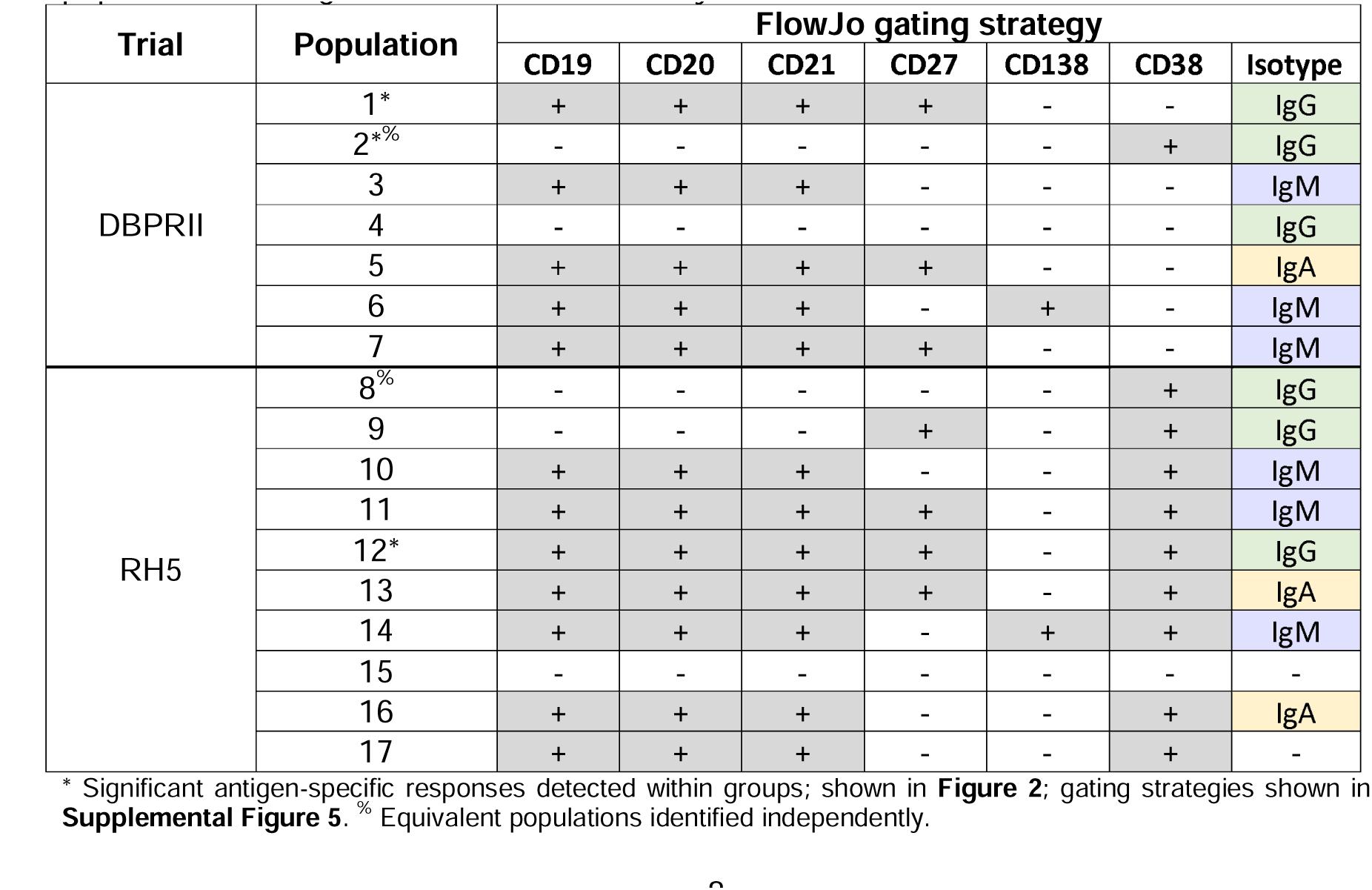
Main peripheral B cell populations as agnostically defined using CITRUS. NB population ordering and numbers are arbitrary.

| Trial | Population | FlowJo gating strategy |  |  |  |  |  |  |
| --- | --- | --- | --- | --- | --- | --- | --- | --- |
|  |  | CD19 | CD20 | CD21 | CD27 | CD138 | CD38 | Isotype |
| DBPRII | 1* | + | + | + | + | - | - | IgG |
|  | 2*% | - | - | - | - | - | + | IgG |
|  | 3 | + | + | + | - | - | - | IgM |
|  | 4 | - | - | - | - | - | - | IgG |
|  | 5 | + | + | + | + | - | - | IgA |
|  | 6 | + | + | + | - | + | - | IgM |
|  | 7 | + | + | + | + | - | - | IgM |
| RH5 | 8% | - | - | - | - | - | + | IgG |
|  | 9 | - | - | - | + | - | + | IgG |
|  | 10 | + | + | + | - | - | + | IgM |
|  | 11 | + | + | + | + | - | + | IgM |
|  | 12* | + | + | + | + | - | + | IgG |
|  | 13 | + | + | + | + | - | + | IgA |
|  | 14 | + | + | + | - | + | + | IgM |
|  | 15 | - | - | - | - | - | - | - |
|  | 16 | + | + | + | - | - | + | IgA |
|  | 17 | + | + | + | - | - | + | - |

\* Significant antigen-specific responses detected within groups; shown in **Figure 2**; gating strategies shown in **Supplemental Figure 5**. % Equivalent populations identified independently.

In the DBPRII trial clusters, significant differences were observed between monthly and delayed dosing in “Population 1” (CD19+CD20+CD21+CD27+CD138-CD38-IgM-IgA-IgG+; **Figure 2B**; **Supplemental Figure 5**) and trends between monthly and delayed or delayed/booster (PA-DB) dosing in “Population 2” (CD19-CD20-CD21-CD27-CD138-CD38+IgM-IgA-IgG+; **Figure 2C**; **Supplemental Figure 5**). While “Population 1” is similar to the CD19+CD21+CD27+IgG+ resting memory population analysed above (**Supplemental Figure 2**), Population 2 was not included in the previous analysis. Responses within other clusters and frequencies of each cluster within total live (B cell-enriched) lymphocytes are shown in **Supplemental Figure 6**.

In the new RH5 trial populations, significant responses were observed in Population 12 (**Figure 2E**; **Supplemental Figure 5**; CD19+CD20+CD21+CD27+CD138-CD38+IgM-IgA-IgG+; again, a similar population to CD19+CD27+IgG+ in **Figure 1F**) in both monthly and delayed booster dosing, with higher responses in delayed boosting vaccinees. Significant differences in post-vaccination RH5-specific responses were not detected between groups in the remaining populations (**Supplemental Figure 7**). No significant responses were observed within Population 8 (CD19-CD20-CD21-CD27-CD138-CD38+IgM-IgA-IgG+; equivalent Population 2 in the DBPRII trial) in the delayed boosting vaccinees.

### DBPRII-specific serum antibody declines more slowly in delayed booster dosing vaccinees

We have previously published ELISA data on serum anti-DBPII total IgG (against the Sal I strain), with an emphasis on comparison of FV+14 responses. Here we observed significantly higher titres with the delayed protein/adjuvant dosing regimen as compared to viral vectors [11]. Now, we extend these analyses to compare the impact of platform and regimen on different isotypes/ subclasses, (**Figure 3A–F**; **Supplemental Figure 8**), durability of serum antibody (**Figure 3G–H**), and immunodominance of subdomain 3 (sd3; **Supplemental Figure 8**). The isotype and subclass analyses showed IgG1, IgG3, IgA and IgA1 responses in both platforms, while IgG4 was detectable solely in the protein/adjuvant vaccinees. No statistically significant post-vaccination IgM responses were observed within individual groups (**Supplemental Figure 8**), while no detectable IgG2 or IgA2 was observed in any sample (not shown). The protein/adjuvant platform also induced higher IgG1, IgG3, IgA and IgA1 (and IgG4) responses as compared to viral vectors (**Supplemental Figure 8A–F**). Within the protein/adjuvant platform, the DBPRII-specific IgG1, IgG4 and IgA1 response was significantly higher in delayed dosing vaccinees (**Figure 3A, C, E**), but comparable between regimens for IgG3 and IgA (**Figure 3B, D**). Median IgM responses were higher in monthly dosing but not statistically significant (**Figure 3F**). Interestingly, we also observed increased anti-DBPRII serum total IgG durability in delayed dosing regimens with both vaccine platforms; a significantly higher fold-change in total serum IgG was observed > 3-months following the peak response in delayed booster vaccinees (VV-D, PA-D; median = 0.45) as compared to monthly dosing vaccinees (VV-M, PA-M; median = 0.15; **Figure 3G**). Enhanced serum durability was also observed with IgG1 in the isotype and subclass analyses (**Figure 3H**). Finally, delayed dosing with PvDBPII appears to have no impact on the immunodominance of the sd3 region (**Supplemental Figure 8**). Equivalent serological analyses from the RH5.1/Matrix-M™ trial will be the focus of a separate report (Silk et al, *manuscript in preparation*).

**Figure 3.**
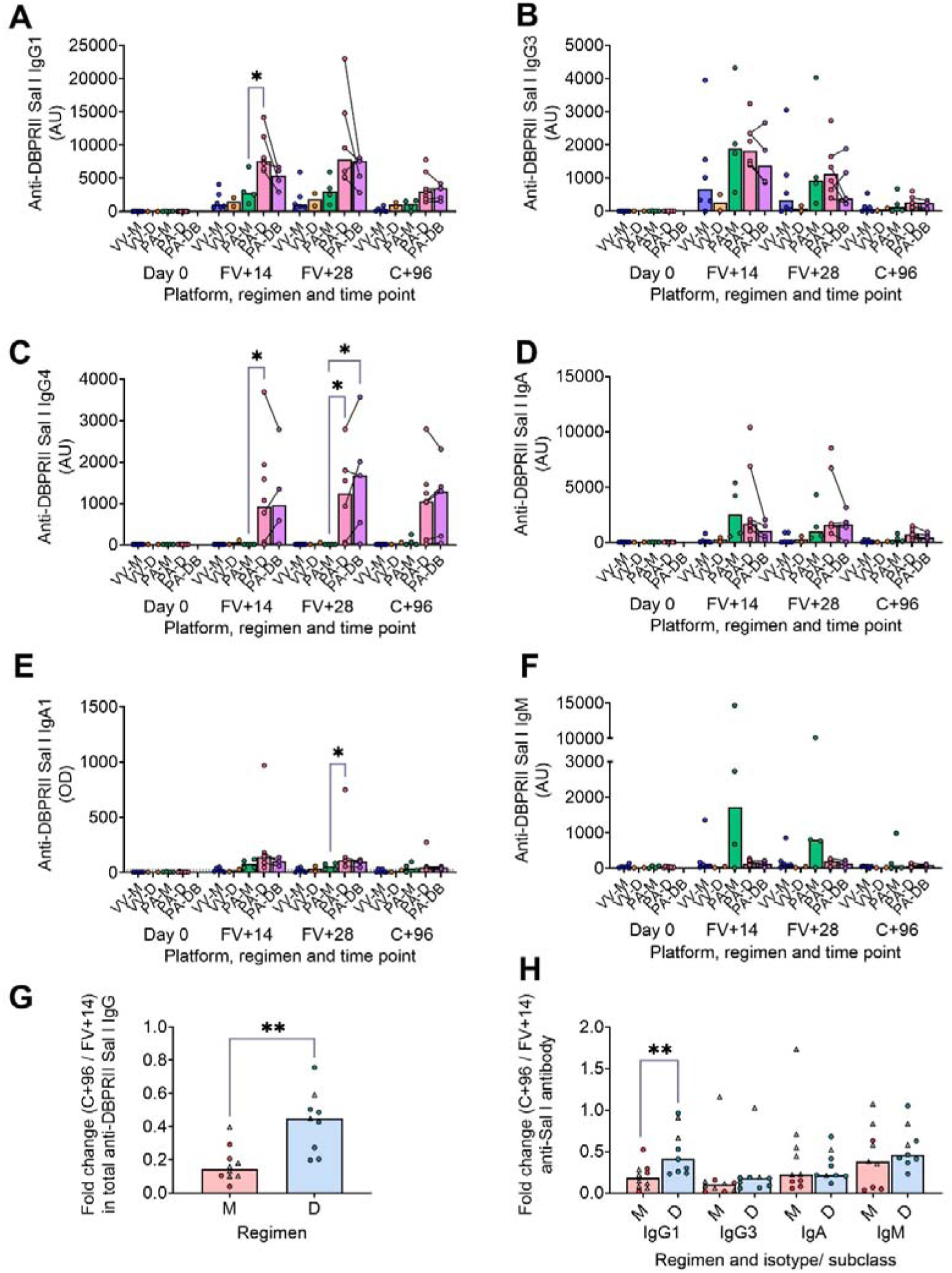
DBPRII-specific peak antibody responses and serum maintenance. Standardised ELISAs were developed to report anti-DBPRII specific antibody responses against the Sal I strain in pre-vaccination (Day 0) and post-final vaccination (FV) serum samples. Responses were compared between protein/adjuvant dosing regimens for IgG1 (**A**), IgG3 (**B**), IgG4 (**C**), IgA (**D**), IgA1 (**E**), and IgM (**F**). Fold change between C+96 and FV+14 was calculated for total IgG (**G**) and specific isotypes/ subclasses (**H**) to compare monthly (M: VV-M, PA-M) and delayed (D: VV-D, PA-D) booster regimens. IgG4 and IgA1 were excluded from this analysis as ≥1 vaccinee had undetectable antibody at both time points. Comparisons between vaccine platforms are shown in **Supplemental Figure 8**. VV-M = ChAd63-MVA viral vector monthly dosing; VV-D ChAd63-MVA delayed booster dosing; PA-M = protein/adjuvant monthly dosing; PA-D = protein/adjuvant delayed booster dosing; PA-DB = protein/adjuvant delayed booster dosing with extra booster. C+96 = 96 days after controlled human malaria infection (approximately 16-weeks after FV). Post-vaccination comparisons were performed between protein/adjuvant dosing regimens by Kruskal Wallis test with Dunn’s correction for multiple comparisons (**A-F**) or fold changes with Mann-Whitney U tests (**G-H**). Sample sizes for all assays were based on sample availability; each circle represents a single sample [triangles indicate viral vector samples in **G-H**]. (**A-F**) VV-M/VV-D/PA-M/PA-D/PA-DB: Day 0 = 6/2/4/8/na, FV+14 = 6/2/4/8/4, FV+28 = 6/2/4/6/5. (**G-H**) M = 9-10, D = 9. Bars represent medians. * *p* < 0.05, ** *p* < 0.01.

### Delayed booster dosing does not impact DBPRII-specific T cell responses

In our main trial report, we showed higher frequencies of IFN-γ-producing effector memory (CD45RA-CCR7-) CD4+ T cells at FV+14 in viral vector as compared to protein/adjuvant vaccinees, with no significant differences observed between monthly and delayed dosing regimens with the latter platform [11]. Here, we extended these analyses to include further time points as well as IL-2/ TNF-α/ IL-5/ IL-13 intracellular cytokine detection (in addition to IFN-γ), allowing a more nuanced comparison of responses between vaccine platform and regimen. Looking first at all DBPRII-specific effector memory CD4+ T cells – based on secretion of any cytokine following stimulation with the DBPRII peptide pool (**Table 3**) – we observed significantly higher frequencies in protein/adjuvant vaccinees at FV+7 as compared to viral vector vaccinees, but no difference between dosing regimens (**Figure 4A–B**). Within the same effector memory CD4+ T cell population, we next assessed total Th1 (defined as IFN-γ and/or IL-2 and/or TNF-α) and Th2 responses (IL-5 and/or IL-13). Both platforms induced Th1 cytokine production and there were no significant differences between platforms or dosing regimens (**Figure 4C-D**). In contrast, the protein/adjuvant vaccines drove higher frequency Th2 responses, comparable across different dosing regimens (**Figure 4E–F**). Trends were similar for both IL-5 and IL-13, although the magnitude of DBPRII-specific IL-5-producing cells was higher (median [range] % DBPRII-specific cells within effector memory CD4+ T cells at FV+7: VV IL-5 = 0.04% [0.00-0.10%], PA IL-5 = 0.60% [0.06-1.35%]; IL-13 VV 0.02% [0.00-0.06%], PA IL-13 = 0.06% [0.02-0.42%]).

**Table 3.**
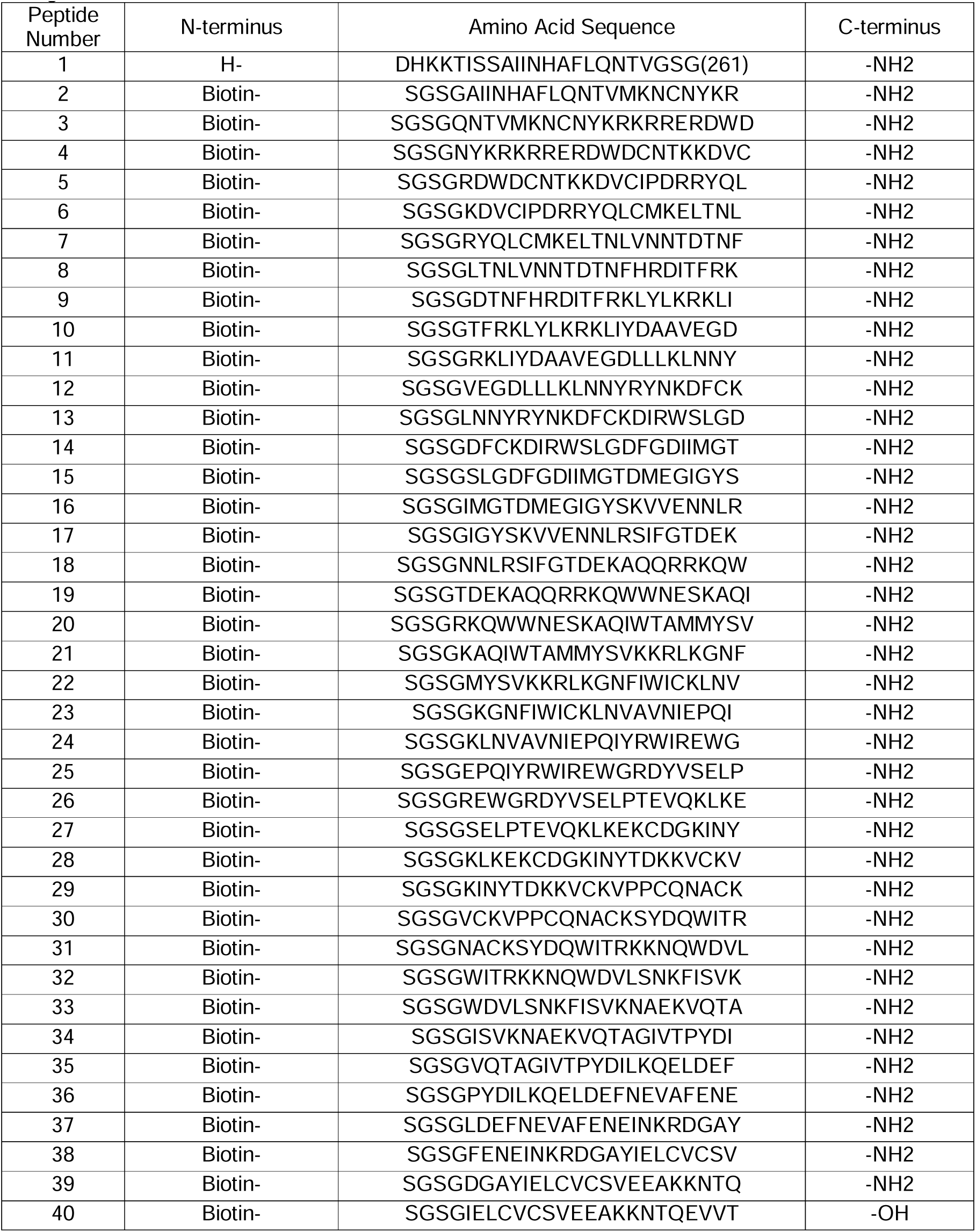
Peptide pool for T cell stimulation. The PvDBPII SalI amino acid sequence was used to design 20mer peptides overlapping by 12 amino acids and these were synthesized by Mimotopes, Australia. Each stock was reconstituted to 50mg/mL in DMSO. A 200µg/peptide/mL working stock of PvDBPII peptides was prepared by adding an equal amount of each peptide to cell culture medium for a final total peptide concentration of 8mg/mL.

**Figure 4.**
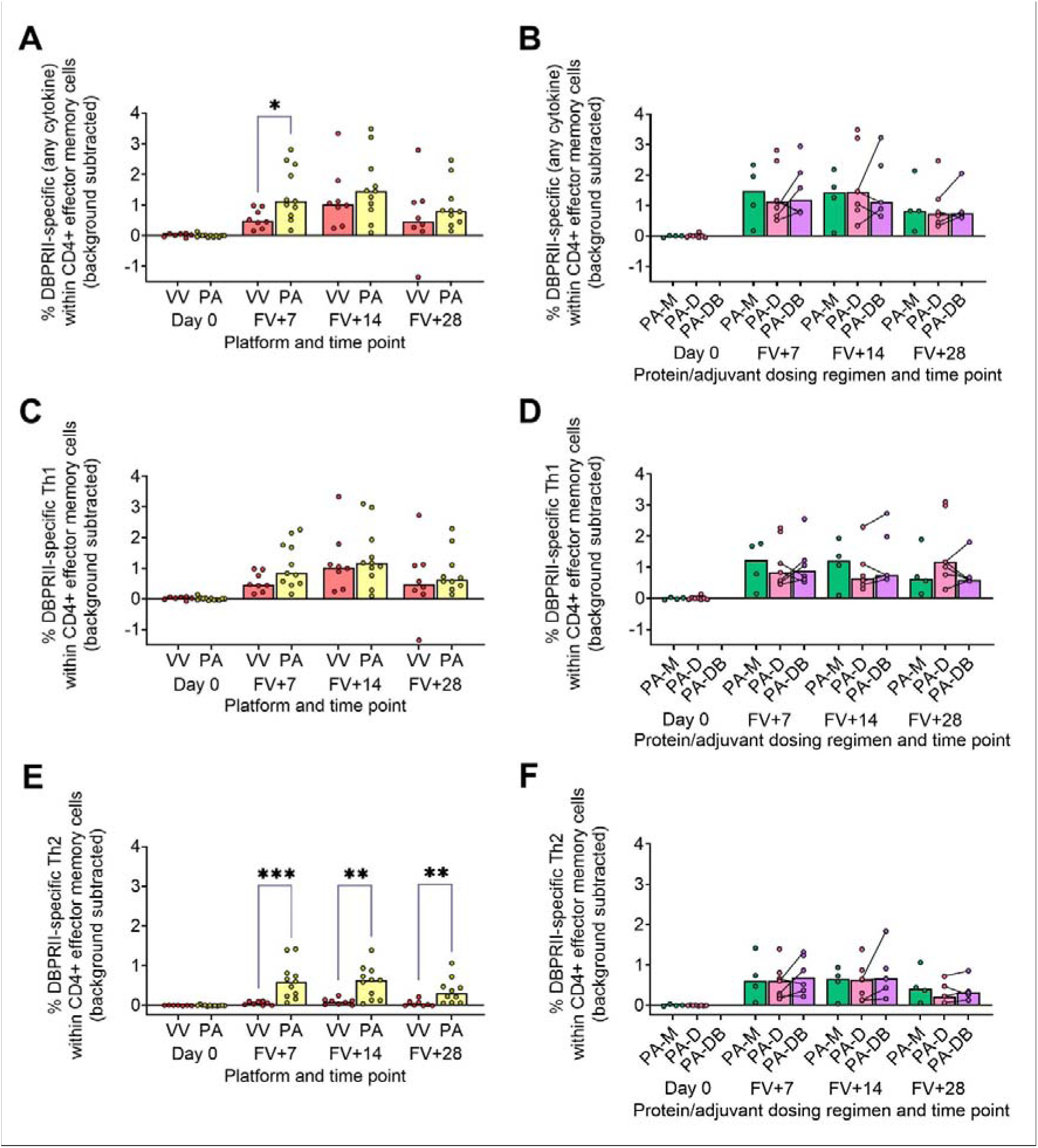
DBPRII-specific CD4+ effector memory T cell responses. PBMC from pre-vaccination (Day 0) and post-final vaccination (FV) time points were analysed for T cell responses by intracellular cytokine staining; gating strategies are as described in Methods and **Supplemental Figure 9**. In brief, DBPRII-specific effector memory CD4+ T cells are reported as frequencies producing cytokines in response to peptide stimulation after background subtraction of cytokine-positive cells in matched samples cultured with media alone. DBPRII-specific responses were compared between vaccine platforms or protein/adjuvant dosing regimens as defined by production of any cytokine [IL-2, IL-5, IL-13, IFN-γ, TNF-α] (**A-B**), any Th1 cytokine [IL-2, IFN-γ, TNF-α] (**C-D**), or any Th2 cytokine [IL-5, IL-13] (**E-F**). CD8+ effector memory T cell responses are shown in **Supplemental Figure 9**. VV = ChAd63-MVA viral vectors; PA = PvDBPII protein/adjuvant [PA-M and PA-D]; PA-M = PvDBPII protein/adjuvant monthly dosing; PA-D = PvDBPII protein/adjuvant delayed booster dosing; PA-DB = PvDBPII protein/adjuvant delayed booster dosing with extra booster. Post-vaccination comparisons were performed between PvDBPII platforms by Mann Whitney U test (**A, C, E**), or protein/adjuvant dosing regimens by Kruskal Wallis test with Dunn’s correction for multiple comparisons (**B, D, F**). Sample sizes for all assays were based on sample availability; each circle represents a single sample. (**A, C, E**) VV/PA: Day 0 = 7/12, FV+7 = 8/11, FV+14 = 8/11, FV+28 = 8/10. (**B, D, E**) PA-M/PA-D/PA-DB: Day 0 = 4/8/na, FV+7 = 4/7/6, FV+14 = 4/7/5, FV+28 = 4/6/5. PA-D vaccinees returning in the PA-DB group are connected by lines. Bars represent medians. * *p* < 0.05, ** *p* < 0.01, *** *p* < 0.001.

Finally, we similarly quantified the effector memory CD8+ T cell response. Here, higher IFN-γ responses were observed with the viral vector platform; no significant responses were observed within the protein/adjuvant platform (**Supplemental Figure 9**).

### DBPRII-specific B cell responses correlate with *in vivo* efficacy against *P. vivax* parasites and durability of anti-DBPRII serum IgG

To confirm the biological relevance of increased B cell immunogenicity in delayed dosing vaccinees we performed Spearman correlations with *in vivo* growth inhibition of blood-stage parasites following *P. vivax* CHMI [11]. The magnitude of the circulating DBPRII-specific memory IgG+ B cell response correlated strongly with IVGI (**Figure 5A**), as did the frequency of DBPRII-specific plasma cells (**Figure 5B**). Since IVGI is calculated over a relatively short time frame following parasite inoculation, we were also interested in assessing the relationship between these B cell responses and the fold change in serum antibody between the peak (FV+14) and a later time point (C+96, approximately 16-weeks later; **Figure 3G**). DBPRII-specific IgG+ memory B cells again correlated strongly with durable serum anti-DBPRII IgG responses (**Figure 5C**), while no association was observed with peak FV+7 plasma cells (**Figure 5D**). Neither IVGI nor serum antibody maintenance correlated with DBPRII-specific Th2 effector memory CD4+ T cell responses (data not shown), suggesting a lesser role for T cell immunogenicity in DBPRII vaccine-mediated protection.

**Figure 5.**
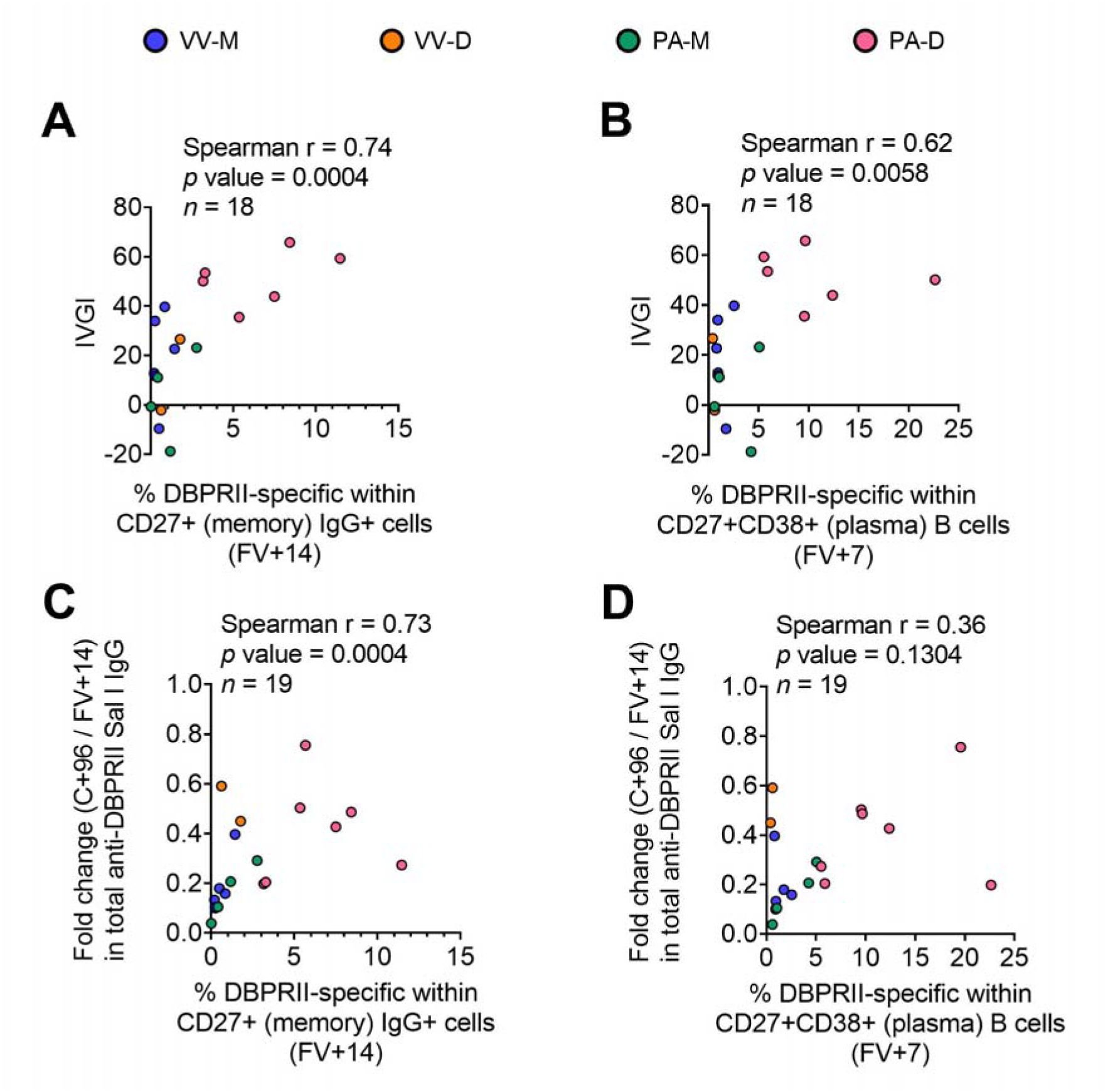
Correlations between circulating DBPRII-specific B cells and *in vivo* growth inhibition of *P. vivax* parasites or maintenance of serum antibody. *In vivo* growth inhibition (IVGI) of *P. vivax* parasites following post-vaccination controlled human malaria infection (CHMI) was calculated from qPCR data as described in the Methods. Spearman correlations were performed between IVGI and the peak frequency of DBPRII-specific memory IgG+ B cells at FV+14 (**A**) or plasma cells at FV+7 (**B**) as defined in **Supplemental Figure 1** and reported in Figure 1. Spearman correlations were also performed between C+96/FV+14 fold change in total anti-DBPRII IgG (Sal I strain; see Figure 3) and memory IgG+ B cells at FV+14 (**C**) or plasma cell at FV+28 (**D**). VV-M = ChAd63-MVA viral vector monthly dosing; VV-D ChAd63-MVA delayed booster dosing; PA-M = protein/adjuvant monthly dosing; PA-D = protein/adjuvant delayed booster dosing. C+96 = 96 days after controlled human malaria infection (approximately 16-weeks after FV). Spearman rho, *p* values and sample sizes are annotated on individual graphs. Each circle represents a single sample.

### Bone marrow plasma cells correlate with serum antibody and circulating memory B cells in adult RH5.1/Matrix-M™ vaccinees

While bone marrow aspirates were not taken in the DBPRII clinical trials, we had the rare opportunity to investigate bone marrow plasma cells with the RH5.1/Matrix-M™ adult vaccinees – the first bone marrow analyses in the context of malaria vaccination [13]. Here, we observed a trend to higher frequencies of RH5-specific cells within total bone marrow B cells in the delayed dosing regimen (**Figure 6A**) which correlated strongly with matched time point serum anti-RH5 IgG (**Figure 6B**). We were also interested to see if the frequencies of RH5-specific cells within circulating B cell populations of interest correlated with vaccine-specific seeding of the bone marrow. We therefore performed additional Spearman correlation analyses between the frequency of RH5-specific cells within “Population 12” (CD19+CD20+CD21+CD27+ CD138-CD38+IgA-IgM-IgG+; the B cell population with greatest differences between regimens) at both matched time points (i.e. V1 or FV+28; **Figure 6C**) or with FV+14 (**Figure 6D**). We observe strong correlations between RH5-specific circulating memory B cells and bone marrow plasma cells at both time points. Similar results were obtained if correlations were performed with RH5-specific CD19+CD27+IgG+ B cells (**Figure 1F**; data not shown). RH5-specific bone marrow B cells did not correlate with circulating RH5-specific CD27+CD38+ (plasma) cells at FV+7 (data not shown).

**Figure 6.**
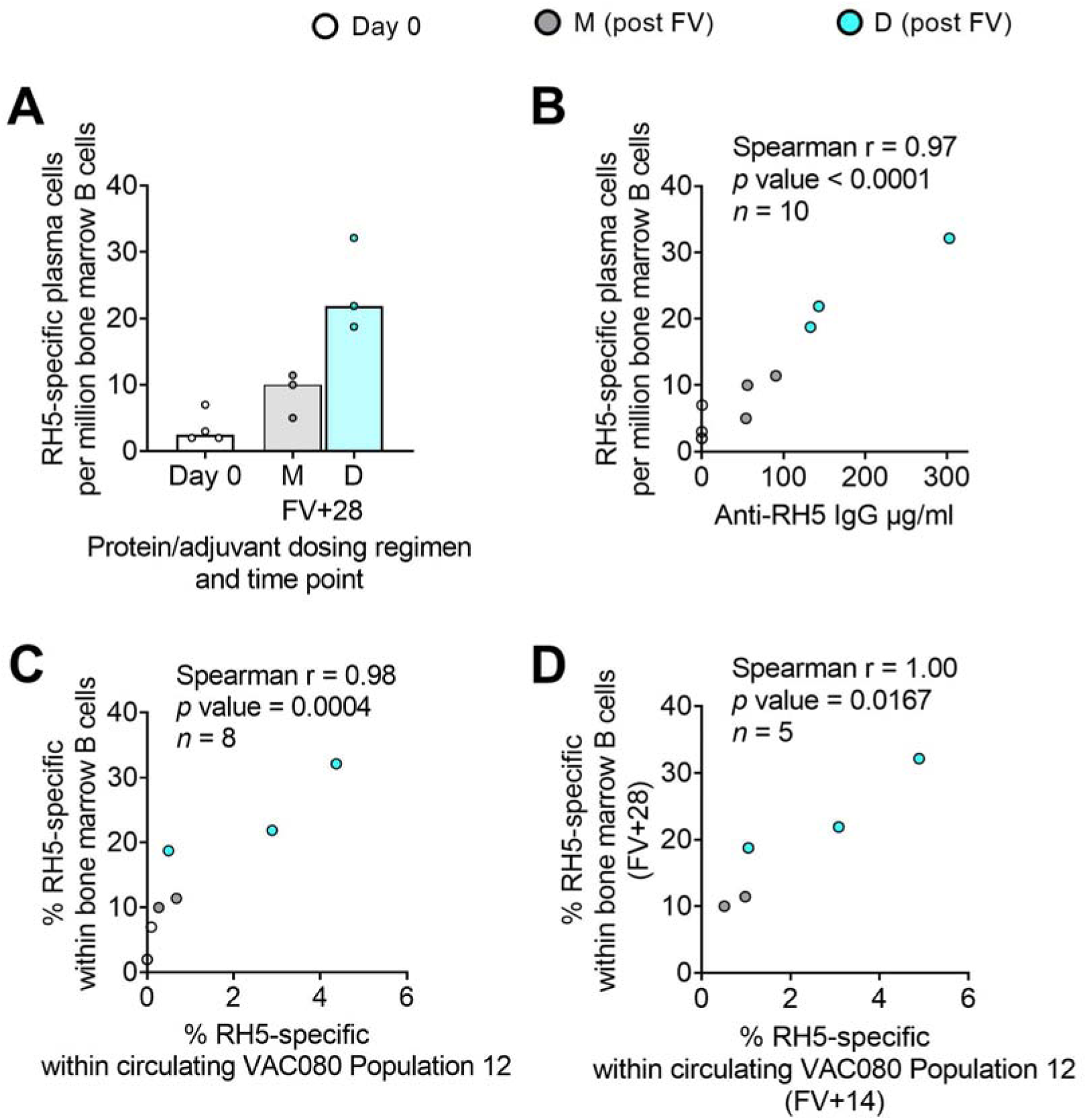
RH5-specific bone marrow plasma cell responses and correlations with serum antibody or circulating RH5-specific cells. RH5-specific bone marrow plasma cells were detected in B cells enriched from pre-and post-final vaccination (FV) bone marrow mononuclear cells and assayed by IgG antibody-secreting cell ELISPOT as described in the Methods. The frequency of RH5-specific IgG plasma cell (antibody-secreting cell) per million bone marrow B cells was compared between dosing regimens (**A**). Spearman correlation analyses were performed between RH5-specific bone marrow B cells and matched time point serum IgG (**B**), matched time point frequency of RH5-specific cells within CITRUS-guided “Population 12” [CD19+CD20+CD21+CD27+CD138-CD38+IgM-IgA-IgG+; see **Table 2**, Figure 2, **Supplemental Figure 5**] (**C**), and between RH5-specific bone marrow B cells at FV+28 and Population 12 at FV+14 (**D**). M = RH5.1/adjuvant monthly dosing; D = RH5.1/adjuvant delayed booster dosing. Post-vaccination comparisons were performed between dosing regimens by Mann Whitney U test (**A**; not significant). Spearman rho, p values and sample sizes are annotated on individual graphs. Each circle represents a single sample.

## Discussion

The data presented here contribute to growing efforts seeking to understand the impact of modifiable vaccine delivery parameters – i.e. vaccine platform or dosing regimen – on protective immune responses. Using samples from independent clinical trials with the blood-stage malaria antigens DBPRII and RH5, we observe a hierarchy of serum antibody and B cell immunogenicity from heterologous viral vectors to monthly protein/adjuvant booster dosing to delayed booster protein/adjuvant vaccination. Consistent with a role for vaccine-specific B cells in sustained humoral immunity, circulating plasma or memory B cell responses correlate with available *in vivo* growth inhibition (IVGI) and durable serum IgG data from the DBPRII trial. In fact, DBPRII-specific memory IgG+ B cells correlate more strongly with IVGI than our previously reported associations between IVGI and serum anti-DBPRII IgG, DARC (the DBP ligand) binding inhibition, or *in vitro* growth inhibitory activity (GIA) with a transgenic *P. knowlesi* strain expressing *P. vivax* DBPRII [11]). In contrast, while differences in CD4+ and CD8+ T cell responses are observed between platforms by intracellular cytokine staining (ICS), there are no discernible differences between protein/adjuvant dosing regimens and no correlation is observed between IVGI and T cell immunogenicity [11].

Our conclusions are in line with data published by Payne *et al*. showing an increase in SARS-CoV-2 spike-specific IgG+ antibody-secreting B cells by ELISPOT 4-weeks after the second dose with longer intervals between BNT162b2 mRNA doses in naïve vaccinees (median = 3.4 weeks, versus median = 10.1 weeks). More mixed results were observed with T cell immunogenicity. For example, the authors observed decreased spike-specific IFN-γ ELISPOT responses and CD8+ IFN-γ by ICS, alongside increased spike-specific CD4+ IL-2 and IFN-γ by ICS [3]). Likewise, recent work from Nicolas *et al.* reported “long interval” (i.e. delayed) booster dosing increases circulating SARS-CoV-2 RBD-specific IgG+ B cells 1-3 weeks after the second mRNA dose, without driving major differences in memory CD4+ or CD8+ T cells (as measured by activation-induced marker [AIM] or ICS [14]). Our own previously published work with a similar AIM assay in the context of a UK RH5.1/AS01 Phase I vaccine trial showed no effect of delayed boosting on the magnitude of the Tfh cell response, but we did detect a slight shift towards a Tfh2 phenotype as compared to monthly boosting vaccinees [4]. To note, the Nicolas *et al*. study compared “short” (median = 3.0 weeks) and “long” (median = 15.8 weeks) intervals between the two mRNA doses similar to the spacing between 2^nd^ and 3^rd^ doses in the RH5 and CSP (RTS,S) trials. However, for many comparisons of parameters between the malaria and SARS-CoV-2 fields it is important to remember that the “delay” is often less substantial in SARS-CoV-2 trials (i.e. a few weeks [1; 2; 3], rather than several months as tested with DBPRII/ RH5/ CSP-based vaccines [4; 5; 6; 7; 8; 9; 11]). We are far from understanding the optimal spacing of booster doses, but if the benefit to B cell immunogenicity with delayed booster vaccination relates to allowing circulating antibody and/or ongoing germinal centres to wane – as we have previously proposed [5] – then it seems likely that this difference of weeks versus months will be immunologically relevant.

Our approach to the B cell flow cytometry analyses also incorporated an agnostic approach to identifying the main circulating B cells populations through use of the CITRUS clustering tool. Significant post-vaccination responses were observed within DBPRII “Population 1” and RH5 “Population 12”, which appear to be subsets of the resting memory populations. Interestingly, within the DBPRII trial, substantial post-vaccination responses within the protein/adjuvant delayed boosting groups were detected in a new “Population 2” (CD19-CD20-CD21-CD27-CD138-CD38+IgM-IgA-IgG+). This was an unexpected observation that would have been missed with traditional B cell analytical approaches that start from the premise that all B cells are CD19+. Given our flow cytometry assay includes a negative pan B cell enrichment step and IgG expression is not expected on other lymphocytes, it is likely that the vast majority of these CD38+IgG+ cells are true B cells. At present, there appears to be limited and conflicting published data on similar (healthy) human vaccine-specific CD19-B cells in circulation. For example, Arumugakani *et al*. have reported on influenza-specific IgG-secreting CD19-CD20-CD38hi cells by ELISPOT and concluded this population was at the plasmablast to mature plasma cell transition, but in contrast to our “Population 2” data this population was also CD27hi [15]. Conversely, Mei *et al*. did not detect post-vaccination tetanus-toxoid specific cells within circulating CD19-CD38+IgG+ B cells by intracellular probe staining and concluded plasma cell CD19 downregulation does not occur until *in situ* in the bone marrow [16].

In light of the interest in CD19-B cells in the context of long-lived plasma cell responses future trials with larger sample sizes should include immunokinetic investigations of CD19-subpopulations and their biological significance. Indeed, of great interest is the strong correlation detected between circulating serum IgG or memory B cells and vaccine-specific bone marrow plasma cells in the RH5 trial. Very few vaccine studies have included lymphoid tissue sampling (reviewed in [13]) and, to the best of our knowledge, this trial represents the first direct analysis of human bone marrow-resident plasma cells in the context of malaria. Since long-term serum antibody is maintained through secretion by long-lived plasma cells in the bone marrow, understanding the factors that impact this compartment is central to optimising durability of humoral immunity. Our data strongly suggest that delayed dosing improves seeding of bone marrow plasma cells, as compared to monthly booster dosing. Future studies should build on these exciting findings with larger sample sizes or sample volumes, which would permit more detailed analyses of populations of interest such as the putative long-lived plasma cell population (CD19-CD38+CD138+ [16; 17; 18]) within total bone marrow plasma cells.

There are also several aspects of the DBPRII serology data that deserve further comment. Firstly, it is interesting to note that median peak responses after an additional booster vaccination (PA-DB vs PA-D) are lower across the majority of isotypes and subclasses measured. This is mirrored in the DBPRII-specific B cell data and indeed reaches statistical significance for the CD27+CD38+ plasma cell response. The exception is serum IgG4 which trends to a higher peak concentration following PA-DB as compared to the PA-D regimen, and in fact we have previously observed an enhanced IgG4 response with higher antigen doses of RH5.1/AS01 in an equivalent UK population [5]. Secondly, we were surprised by the absence of detectable IgG2 following vaccination with any of the platform/ regimens. This is in contrast to the previous RH5 analyses where we observed a (low) IgG2 response to both monthly and delayed booster dosing regimens, with better serum maintenance in the latter group [5]. Finally, while intragroup variation and small sample sizes reduced the statistical power to detect differences in the IgM analyses, it is interesting to note that median responses were higher in the monthly as compared to delayed dosing regimens (FV+14: PA-M = 1708 AU, PA-D = 119.8 AU, PA-DB = 126.0 AU; FV+28: PA-M = 794.1 AU, PA-D = 204.4 AU, PA-DB = 129.4 AU). Although frequencies of DBPRII-specific B cells within the memory IgM+ population were very low, these did trend to slightly higher medians at FV+28 and correlate with serum IgM (Spearman r = 0.69, *p* = 0.0014, *n* = 18).

Given the multitude of parameters assayed, machine learning – such as with the SIMON platform [19] – would have been a useful strategy for interrogating which read-out or set of read-outs best predicted our trial outcomes of interest e.g. peak vaccine-specific IgG, IVGI (DBPRII trial only) or seeding of bone marrow plasma cells (RH5 trial only). Unfortunately, this approach was precluded by insufficient sample size. Indeed, the small sample sizes of the different vaccination groups represents the main limitation of our analyses for both trials.

To conclude, our data indicate that while changing vaccine platform drives broad effects on post-vaccination immune responses, modulating booster dosing regimen more narrowly impacts humoral immunity. Importantly, these differences in immunogenicity appear to have relevance for protection from *P. vivax* in a CHMI model. While this investigation of the delayed booster regimen was a serendipitous effect of the SARS-CoV-2 pandemic – not unlike the original delayed fractional booster observations with RTS,S [6]– it now seems likely that future clinical development of the PvDBPII candidate will benefit from further interrogation of delayed booster regimens. These findings are supported by data from an independent RH5.1/Matrix-M™ clinical trial in malaria-exposed adults in Tanzania where delayed booster dosing not only increases the frequency of circulating RH5-specific memory B cells, but also RH5-specific plasma cells in the bone marrow.

## Methods

### Clinical trials

This study focused on the comparison of immune responses between groups vaccinated with the *Plasmodium vivax* antigen DBPRII with different platforms and dosing regimens (**Table 1** [11]). In brief, two Phase I/IIa vaccine efficacy trials (NCT04009096 and NCT04201431) were conducted in parallel at a single site in the UK (Centre for Clinical Vaccinology and Tropical Medicine, University of Oxford). NCT04009096 was an open label trial to assess the ChAd63 and MVA viral-vectored vaccines encoding PvDBPII (VV-PvDBPII), while the NCT04201431 trial assessed the protein vaccine PvDBPII in Matrix-M™ adjuvant from Novavax (PvDBPII/M-M). NCT04009096 viral vectors were administered at 0, 2 months (VV-M) or in a delayed dosing regimen (0, 17, 19 months; VV-D). For NCT04201431, the protein/adjuvant was administered monthly (0, 1, 2 months; PA-M) or in a delayed dosing regimen (0, 1, 14 months; PA-D). A subset of vaccinees from the protein/adjuvant delayed dosing regimen returned for an additional booster at 19 months (PA-DB). ChAd63-PvDBPII was administered at a dose of 5×10^10^ viral particles, MVA-PvDBPII at 2×10^8^ plaque forming units, and PvDBPII protein at 50μg mixed with 50μg Matrix-M™. Delayed regimens were due to trial halts during the pandemic. Eligible vaccinees were healthy, Duffy-positive, malaria-naïve adults, aged 18 to 45 years. Trials were approved by the UK National Health Service Research Ethics Services (REC; references 19/SC/0193 and 19/SC/0330) as well as by the UK Medicine and Healthcare products Regulatory Agency (MHRA; reference CTA 21584/0414/001-0001 and CTA 21584/0418/001-0001). All vaccinees gave written informed consent.

This study also includes analyses of samples from a further Phase Ib clinical trial with *Plasmodium falciparum* vaccine candidate RH5.1 (50µg) in Matrix-M™ adjuvant (50µg) in malaria-endemic setting in Tanzania (NCT04318002). RH5.1/Matrix-M™ was administered at either a monthly 0, 1, 2 months (M) or delayed 0, 1, 6 months (D) booster regimen. The final vaccination in the delayed booster regimen was given at a fractionated RH5.1 antigen dose of 10µg, rather than 50µg (the dose of Matrix-M™ was not fractionated and remained 50 µg). Eligible vaccinees were healthy adults (negative for malaria by blood smear at screening), aged 18 to 45 years. The trial was approved by the Tanzanian Medicines and Medical Devices Authority (reference TMDA0020/CTR/0006/01), the National Institute for Medical Research (references NIMR/HQ/R.8a/Vol.IX/3537 and NIMR/HQ/R.8c/Vol.1/1887), the Ifakara Health Institute Institutional Review Board (reference IHI/IRB/No:49-2020), and the Oxford Tropical Research Ethics Committee (reference 9-20). All vaccinees gave written informed consent.

## Methods details

### Flow cytometry – B cells

Cryopreserved PBMC from the DBPRII trial were thawed into R10 media (RPMI [R0883, Sigma] supplemented with 10% heat-inactivated FCS [60923, Biosera], 100U/ml penicillin / 0.1mg/mL streptomycin [P0781, Sigma], 2mM L-glutamine [G7513, Sigma]) then washed and rested in R10 for 1h. B cells were enriched (Human Pan-B cell Enrichment Kit [19554, StemCell]) and then stained with viability dye FVS780 (565388, BD Biosciences). Next, B cells were stained with anti-human CD19-BV786 (563325, BD Biosciences), anti-human CD20-BUV395 (563782, BD Biosciences), anti-human IgG-BB515 (564581, BD Biosciences), anti-human IgM-BV605 (562977, BD Biosciences), anti-human CD27-PE-Cy7 (560609, BD Biosciences), anti-human CD21-BV711 (563163, BD Biosciences), anti-human CD38-BV480 (566137, BD Biosciences), anti-human CD138-APC-R700 (566050, BD Biosciences), anti-human IgA-PerCP-Vio700 (130-113-478, Miltenyi) as well as two fluorophore-conjugated DBPRII probes. Preparation of the DBPRII probes was based on our previously published protocols with the *P. falciparum* blood-stage malaria antigen RH5 [12; 20]. In brief, monobiotinylated DBPRII was produced by transient co-transfection of HEK293F cells with a plasmid encoding BirA biotin ligase and a plasmid encoding a monoFC-fused, biotin acceptor peptide-and c-tagged full-length DBPRII. Monobiotinylated DBPRII was purified by affinity chromatography (c-tag) and size exclusion chromatography. The monoFC solubilisation domain was cleaved using TEV protease. Probes were freshly prepared for each experiment, by incubation of monobiotinylated DBPRII with streptavidin-PE (S866, Invitrogen) or streptavidin-APC (Biolegend, 17-4317-82) at an approximately 4:1 molar ratio to facilitate tetramer generation and subsequently centrifuging to remove aggregates. Following surface staining, cells were permeabilised and fixed with Transcription Factor Buffer Set (562574, BD Biosciences), stained with anti-human Ki67-BV650 (563757, BD Biosciences), washed, and stored at 4°C until acquisition. Samples were acquired on a Fortessa X20 flow cytometer with FACSDiva8.0 (both BD Biosciences). Samples were analysed using FlowJo (v10; Treestar). Samples were excluded from analysis if <50 cells in the parent population.

The B cell assay with cryopreserved samples from the RH5 clinical trial was performed as above with two modifications to the protocol. First, RH5 probes rather than DBPRII probes were used as previously described [12; 20]. Second, probe staining was repeated during the intracellular cytokine staining step at a 1/10 dilution as compared to concentrations used for surface staining.

See below for details of CITRUS analyses with B cell flow cytometry samples.

### Flow cytometry – T cells

DBPRII peptide stimulation was used to detect DBPRII-specific T cells in an intracellular cytokine staining (ICS) assay as previously described [11]. Cryopreserved PBMC were thawed in R10 and rested before an 18h stimulation with medium alone, 2.5 µg/peptide/mL of a PvDBPII 20mer peptide pool (Mimotopes; **Table 3**), or 1 µg/mL Staphylococcal enterotoxin B (SEB; S-4881, Sigma; positive control). Anti-CD28 (1µg/ml; 16-0289-85, eBioscience), anti-CD49d (1µg/ml; 16-0499-85, eBioscience) and anti-CD107a-PE-Cy5 (15-1079-42, eBioscience) were included in the cell culture medium. Brefeldin A (00-4506-51, eBioscience) and monensin (00-4505-51, eBioscience) were added after 2h. Following incubation, PBMC were stained with viability dye Live/Dead Aqua (L34966, Invitrogen) and anti-human CCR7-BV711 (353228, Biolegend). Cells were then permeabilised and fixed with Cytofix/Cytoperm (554714, BD Biosciences) before staining with anti-human CD14-eF450 (48-0149-42, eBioscience), anti-human CD19-eFl450 (48-0199-42, eBioscience), anti-human CD8a-APC-eF780 (47-0088-42, eBioscience), anti-human IFN-γ-FITC (11-7319-82, eBioscience), anti-human TNFα-PE-Cy7 (25-7349-8, eBioscience), anti-human CD3-AF700 (56-0038-82, eBioscience), anti-CD4-PerCP Cy5.5 (300530, Biolegend), anti-human IL-2-BV650 (500334, Biolegend), anti-human IL5-PE (500904, Biolegend), anti-human IL13-APC (501907 Biolegend), anti-human CD45RA-BV605 (304134, Biolegend). Finally, cells were washed, and stored at 4°C until acquisition. Samples were acquired on a Fortessa X20 flow cytometer with FACSDiva8.0 (both BD Biosciences). Samples were analysed using FlowJo (v10; Treestar). Background cytokine responses to medium alone were subtracted from DBPRII-specific responses. Samples were excluded from analysis if <50 cells in the parent population.

### ELISAs

For the DBPRII clinical trial, antigen-specific total IgG, IgG3, IgG4, IgA, IgA1 and IgM titres were determined by standardised ELISA in accordance with published methodology [21]. Nunc MaxiSorp™ flat-bottom ELISA plates (44-2404-21, Invitrogen) were coated overnight with 2µg/mL (for total IgG titres) or 5µg/mL (for IgG1, IgG3, IgG4, IgA, IgA1 and IgM titres) of DBPRII SalI protein or 2µg/mL of sd3 protein in PBS. DBPRII protein was produced as previously described [11], while sd3 protein was produced by transient transfection of Expi395F cells with a plasmid encoding a monoFc, DBP Subdomain 3 (sequence as per UniProt P22290 PVDR residues P387-S508) and a C-terminal c-tag. The monoFc was cleaved using TEV protease and sd3 was purified by affinity chromatography (c-tag) and size exclusion chromatography. Plates were washed with washing buffer composed of PBS containing 0.05% TWEEN® 20 (P1379, Sigma-Aldrich) and blocked with 100µL of Starting Block™ T20 (37538, ThermoFisher Scientific). After removing blocking buffer, standard curve and internal controls were diluted in blocking buffer using a pool of high-titre vaccinee plasma or serum, specific for each antigen and isotype or subclass being tested, and 50µL of each dilution was added to the plate in duplicate. Test samples were diluted in blocking buffer to a minimum dilution of 1:50 (or 1:100 for total IgG) and 50µL was added in triplicate. Plates were incubated for 2 hours at 37°C (or 20°C for total IgG) and washed in washing buffer. An alkaline phosphatase-conjugated secondary antibody was diluted at the manufacturer’s recommend minimum dilution for ELISA in blocking buffer. The antibody used was dependent on the isotype or subclass being assayed and were as follows: IgG-AP (A3187, Thermo Scientific), IgG1 Fc-AP (9054-04, Southern Biotech), IgG3 Hinge-AP (9210-04, Southern Biotech), IgG4 Fc-AP (9200-04, Southern Biotech), IgA-AP (2050-04, Southern Biotech), IgA1-AP (9130-04, Southern Biotech), and IgM-AP (2020-04, Southern Biotech). 50µL of the secondary antibody dilution was added to each well of the plate and incubated for 1 h at 37°C (or 20°C for total IgG). Plates were developed using PNPP alkaline phosphatase substrate (N2765, Sigma-Aldrich) for 1-4 h at 37°C (or approximately 15 minutes at 20°C for total IgG). Optical density at 405 nm was measured using an ELx808 absorbance reader (BioTek) until the internal control reached an OD405 of 1. The reciprocal of the internal control dilution giving an OD405 of 1 was used to assign an AU value of the standard. Gen5 ELISA software v3.04 (BioTek) was used to convert the OD405 of test samples into AU values by interpolating from the linear range of the standard curve fitted to a four-parameter logistics model. Any samples with an OD405 below the linear range of the standard curve at the minimum dilution tested were assigned a minimum AU value according to the lower limit of quantification of the assay. For assessment of IgG2 and IgA2 responses, no anti-DBPRII IgG2 or IgA2 samples were available for standard curve generation. Responses were measured on plates coated with 5µg/mL DBPRII. Four wells were also coated with RH5.1 protein for development control wells. Each sample was tested in duplicate with six negative control serum samples and two development control serum samples on the RH5.1 coated wells from a previous RH5.1/AS01 vaccine trial [4]. Secondary antibodies used were IgG2 Fd-AP (9080-04) and IgA2-AP (9140-04). The assay was carried out as above and plates were developed for 2-4 hours at 37°C.

For the RH5 clinical trial, serum antibody levels to full-length RH5 protein (RH5.1) were assessed by standardised ELISA methodology as previously described [4; 22]. In brief, the reciprocal of the test sample dilution giving an optical density of 1.0 at 405nm (OD_405nm_) was used to assign an ELISA unit value of the standard. The standard curve and Gen5 software v3.04 (Agilent) was then used to convert the OD_405nm_ of test samples to arbitrary units (AU). Responses are reported in µg/mL using conversion factor from AU generated by calibration-free concentration analysis (CFCA) as previously reported [22].

### Bone marrow aspirate processing and ELISPOTs

A single 10mL bone marrow aspirate was collected per vaccinee into EDTA in the RH5 trial. Bone marrow mononuclear cells (BMMNC) were purified from aspirates by density centrifugation on Lymphoprep (1114545, Axis Shield) following passage through a 70µm nylon cell strainer (542070, Greiner Bio-One Ltd). BMMNC were cryopreserved for future use in FCS (S1810, Biosera) with 10% DMSO (D2650, Sigma). Samples were subsequently thawed and enriched for B cells using a Pan B Cell Enrichment Kit (19554, Stemcell) for detection of RH5-specific plasma cells with an antibody-secreting cell (ASC) ELISPOT. In brief, enriched bone marrow B cells were aliquoted onto MAIP ELISpot Plates (MAIPS4510, Millipore) coated with 5µg/mL RH5 protein, PBS (negative control), or 50µg/mL polyvalent goat anti-human immunoglobulin (positive control; H1700 Caltag). Plates were incubated for 16-18h at 37°C prior to cell removal and incubation with anti-human IgG conjugated to alkaline phosphatase (γ-chain specific; 401442, Calbiochem). Finally, plates were developed with BCIP/NBT (M0711A, Europa Bioproducts) and read on an AID ELISPOT Plate Reader (AID). To note, for consistency all ASCs are referred to as plasma cells throughout this report.

### *In vivo* growth inhibition (IVGI)

*In vivo* growth inhibition (IVGI) has been reported elsewhere for the DBPRII trial [11]. In brief, IVGI was calculated for each vaccinee as the percentage reduction in parasite multiplication rate (PMR) relative to the mean PMR of the unvaccinated controls. PMR was modelled for each vaccinee based on log_10_ transformed qPCR data of the *P. vivax* 18S ribosomal RNA gene, using a mean of three replicate qPCR results for each vaccinee per time point [11; 23; 24]. Further details of the IVGI/ PMR methodology in this trial can be found in the Supplementary Appendix of the primary trial report [11].

### Quantification and Statistical Analyses

Comparisons were performed between regimens with (two-tailed) Mann-Whitney tests or Kruskal-Wallis test with Dunn’s Correction for multiple comparisons (GraphPad Prism v9). A *p* value of < 0.05 was considered statistically significant.

Specifics of the CITRUS analyses are outlined in further detail below. For all data, relevant statistical tests and sample sizes are specified in figure legends.

### Clustering of B cell flow cytometry data with CITRUS

Raw fcs files with file-internal compensation (i.e. acquisition-defined) from the B cell flow cytometry assays with DBPRII or RH5 trial samples were uploaded separately into Cytobank. Live, single (B cell-enriched) lymphocytes were gated and then analysed with CITRUS using equal event sampling. All fluorophore channels were used for clustering with the exception of the probes and the viability stain. All groups and time points were run in a single CITRUS analysis per trial. Median expression values for each fluorophore for each CITRUS-defined cluster were exported per sample. Average expression values across all samples for each fluorophore were then calculated per cluster to define FlowJo gating strategies. Clusters with shared gating strategies were combined into new populations (**Table 2**) for re-analysis in FlowJo. Ki67 was included in the CITRUS clustering but median expression values did not facilitate defining a dichotomous grating strategy and thus Ki67 was not utilised in the population definitions. Samples were excluded from analysis if <50 cells in the parent population.

## Data Availability

Data produced in the present study are available upon reasonable request to the authors.

## Acknowledgements

We thank the vaccinees and clinical staff for participating in and running the clinical trials essential for this study, especially Fay Nugent, Jee-Sun Cho, Yrene Themistocleous, Thomas Rawlinson, Alison Lawrie, Ian Poulton, Rachel Roberts, Iona Taylor, Sumi Biswas, Julie Furze, Baktash Khozoee, Nicola Greenwood, Saumu Ahmed, Florence Milando, and Neema Balige. We also thank Jenny Reimer and Cecilia Carnot at Novavax for providing the Matrix-M™ adjuvant, Federica Cappuccini for guidance developing the T cell flow cytometry assay, Robert Hedley and Vasiliki Tsioligka from the Sir William Dunn School of Pathology Flow Cytometry Facility for assistance with sample acquisition and training, and the qPCR team on the NCT04009096 and NCT04201431 trials (Duncan Bellamy, Hannah Davies, Francesca Donnellan, Amy Flaxman, Reshma Kailath, Rebecca Makinson, Indra Rudiansyah, and Marta Ulaszewska). CMN was supported by a Sir Henry Wellcome Postdoctoral Fellowship (209200/Z/17/Z). SJD was supported by a Wellcome Trust Senior Fellowship (106917/Z/15/Z) and is a Jenner Investigator. The NCT04009096 trial was funded by the European Union’s Horizon 2020 research and innovation program under grant agreement 733073 for MultiViVax. The NCT04201431 trial was funded by the Wellcome Trust Malaria Infection Study in Thailand (MIST) program [212336/Z/18/Z]. The DBPRII clinical trial was also supported in part by the UK Medical Research Council (MRC) [G1100086] and the NIH Research (NIHR) Oxford Biomedical Research Center (BRC). The views expressed are those of the author(s) and not necessarily those of the NHS, the NIHR or the Department of Health. Development of PvDBPII as a vaccine candidate was supported by grants from the Biotechnology Industry Research Assistance Council (BIRAC), New Delhi and PATH Malaria Vaccine Initiative. MVDP was supported by grants from the Bill and Melinda Gates Foundation and Department of Biotechnology (DBT), Government of India. This work was also supported in part by grants from Agence Nationale de Recherche to CEC (ANR-18-CE15-0026 and ANR 21 CE15-0013-01). CEC is supported by the French Government’s Laboratoire d’Excellence “PARAFRAP” (ANR-11-LABX-0024-PARAFRAP). The NCT04318002 trial was funded by the EDCTP2 programme supported by the European Union (grant number RIA2016V-1649-MMVC). The views and opinions of authors expressed herein do not necessarily state or reflect those of EDCTP. In both NCT04201431 and NCT04318002 trials the Matrix-M™ adjuvant was provided by Novavax.

## Author contributions

CMN led the study. AIO, AMM and SJD were chief, principal or lead investigators on the clinical trials. JRB, SES, CGM, KMM, MMH, AML, WFK, IMM, KM, MB, HD, LK, NE, SR, CMN performed experiments and/or oversaw critical sample processing. JRB, SES, NE and CMN analysed and/or reviewed data. VSC, PM and CEC contributed the PvDBPII vaccine. JRB and CMN wrote the manuscript.

## Supplemental Figures

**Supplemental Figure 1.**
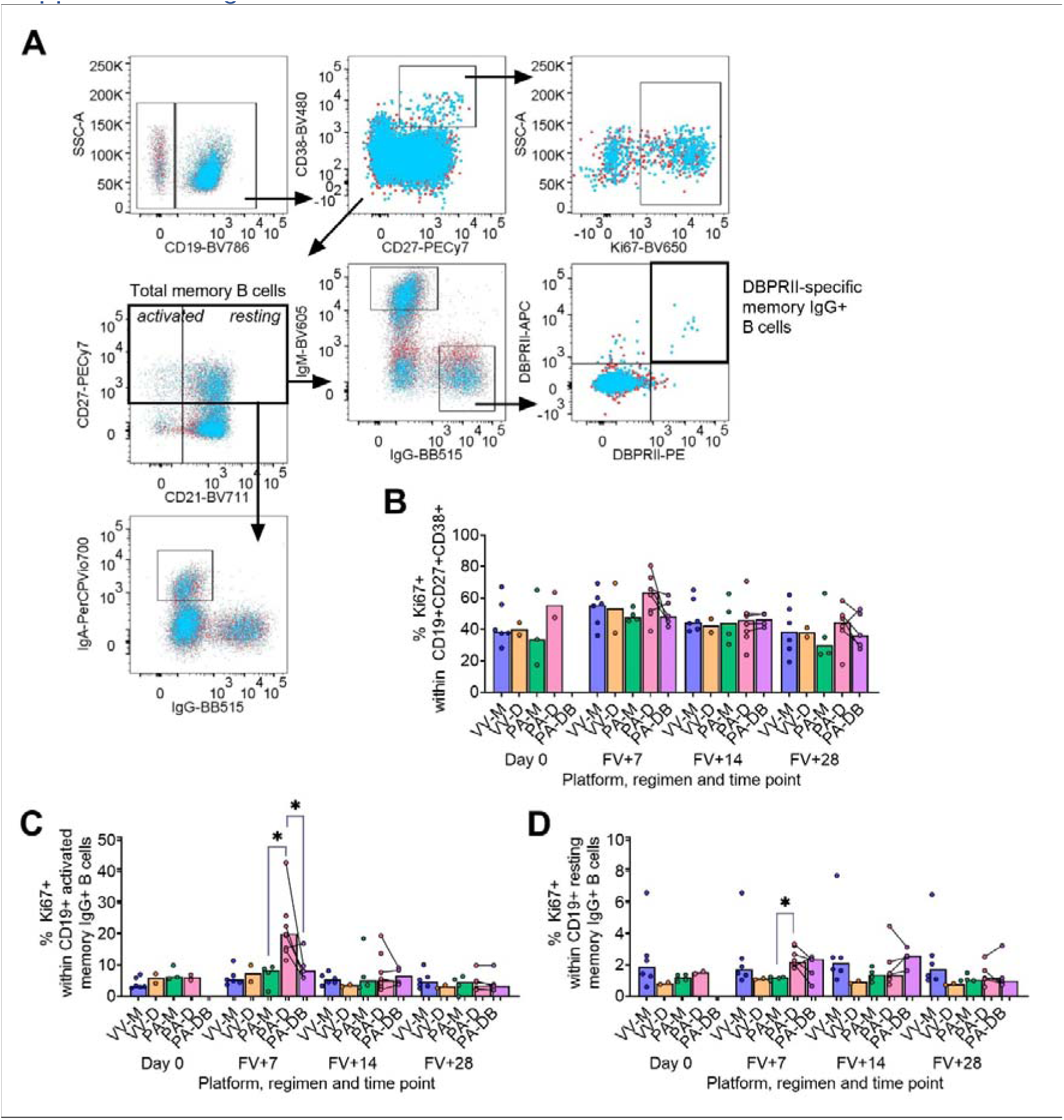
DBPRII B cell gating strategy and expression of proliferation marker Ki67. PBMC from pre-vaccination (Day 0) and post-final vaccination (FV) time points were analysed for B cell responses by flow cytometry. (**A**) Gating strategy shows identification of CD19+ B cells within live single (B cell-enriched) lymphocytes, and definition of CD38+CD27+ plasma cells within this population. Total memory cells are defined as CD27+ non-plasma cells (following use of a NOT gate to exclude plasma cells; indicated with thick black box); activated and resting memory B cells are more specifically categorised as CD21-CD27+ or CD21+CD27+, respectively. IgG+, IgM+ or IgA+ populations are subsequently gated within total memory cells. Proliferating (Ki67+) or vaccine-specific (those co-staining with DBPRII-PE and DBPRII-APC probes as indicated by thick black box) cells are defined within the plasma cell or isotype-specific memory B cell populations. Example shows Ki67 expression of plasma cells and DBPRII-specific gating on memory IgG+ B cells. A FV+14 sample (blue) is overlaid on a matched Day 0 sample (red) for all plots. Frequencies of Ki67+ cells shown within plasma cells (**B**), activated IgG+ memory B cells (**C**), and resting IgG+ memory B cells (**D**). VV-M = ChAd63-MVA viral vector monthly dosing; VV-D ChAd63-MVA delayed booster dosing; PA-M = protein/adjuvant monthly dosing; PA-D = protein/adjuvant delayed booster dosing; PA-DB = protein/adjuvant delayed booster dosing with extra booster. Post-vaccination comparisons were performed between protein/adjuvant dosing regimens by Kruskal Wallis test with Dunn’s correction for multiple comparisons. Sample sizes for all assays were based on sample availability; each circle represents a single sample. VV-M/VV-D/PA-M/PA-D/PA-DB: Day 0 = 6/2/3-4/2/na, FV+7 = 6/2/4/8/5, FV+14 = 6/2/4/8/4, FV+28 = 6/2/4/6/5. PA-D vaccinees returning in the PA-DB group are connected by lines. Bars represent medians. * *p* < 0.05.

**Supplemental Figure 2.**
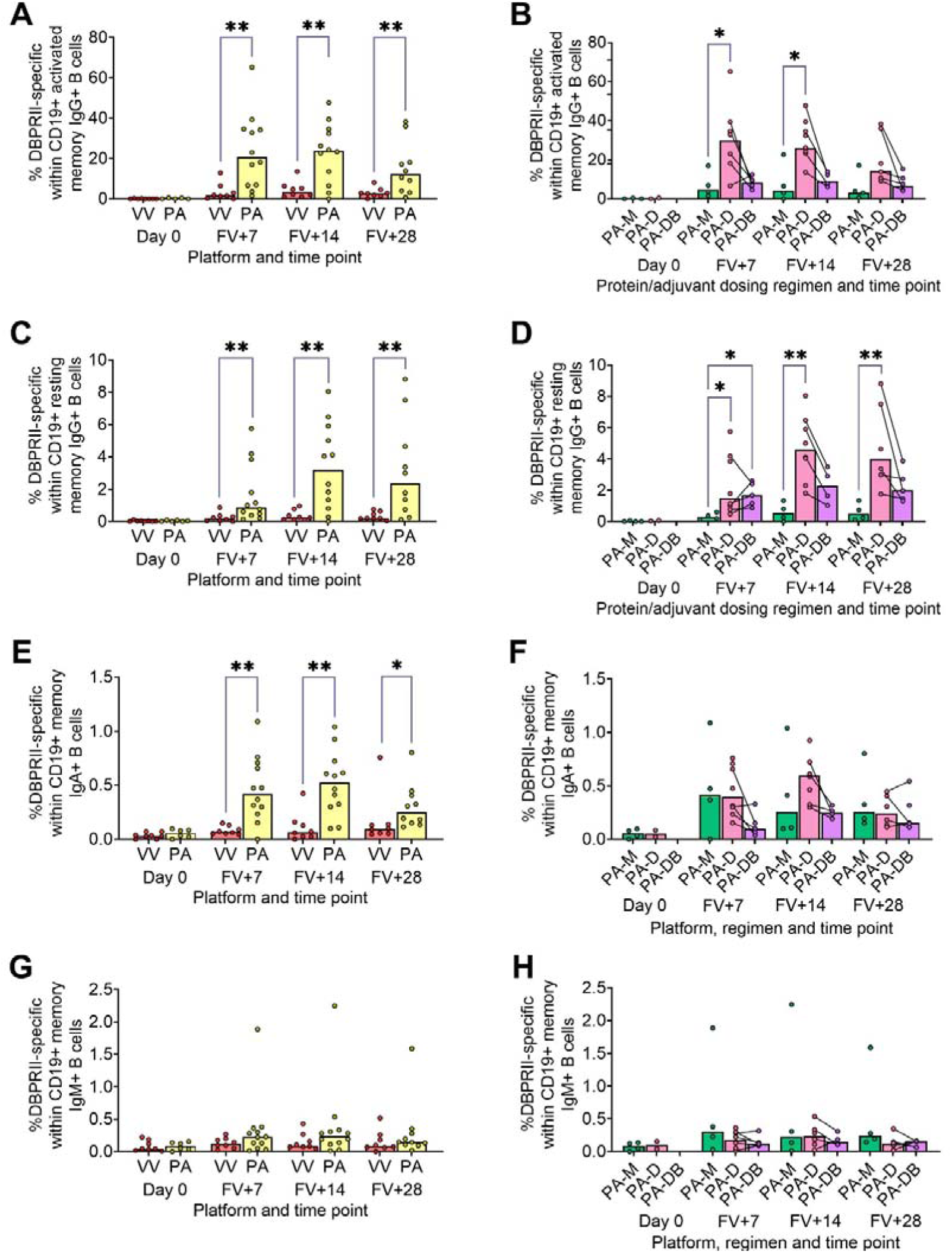
Extended DBPRII-specific memory B cell responses. PBMC from pre-vaccination (Day 0) and post-final vaccination (FV) time points were analysed for B cell responses by flow cytometry; gating strategies are as described in Methods and **Supplemental Figure 1**. Frequencies of DBPRII-specific B cells – identified by probe staining – were compared within activated memory IgG+ B cells (**A-B**), resting memory IgG+ B cells (**C-D**), total memory IgA+ B cells (**E-F**), and total memory IgM+ B cells (**G-H**) between vaccine platforms (**A, C, E, G**) or protein/adjuvant dosing regimens (**B, D, F, H**). VV = ChAd63-MVA viral vectors; PA = PvDBPII protein/adjuvant [PA-M and PA-D]; PA-M = PvDBPII protein/adjuvant monthly dosing; PA-D = PvDBPII protein/adjuvant delayed booster dosing; PA-DB = PvDBPII protein/adjuvant delayed booster dosing with extra booster. Post-vaccination comparisons were performed between DBPRII platforms (**A, C, E, G**) with Mann-Whitney U tests, or between PvDBPII protein/adjuvant dosing regimens by Kruskal Wallis test with Dunn’s correction for multiple comparisons (**B, D, F, H**). Sample sizes for all assays were based on sample availability; each circle represents a single sample. (**A, C, E, G**) VV/PA: Day 0 = 8/5-6, FV+7 = 8/12, FV+14 = 8/12, FV+28 = 8/10. (**B, D, F, H**) PA-M/PA-D/PA-DB: Day 0 = 3-4/2/na, FV+7 = 4/8/5, FV+14 = 4/8/4, FV+28 = 4/6/5. (**E-F**) M/D: Day 0 = 5/1-4, FV+7 = 4-5/3-4, FV+14 = 4/6, FV+28 = 4/4. PA-D vaccinees returning in the PA-DB group are connected by lines. Bars represent medians. * *p* < 0.05, ** *p* < 0.01.

**Supplemental Figure 3.**
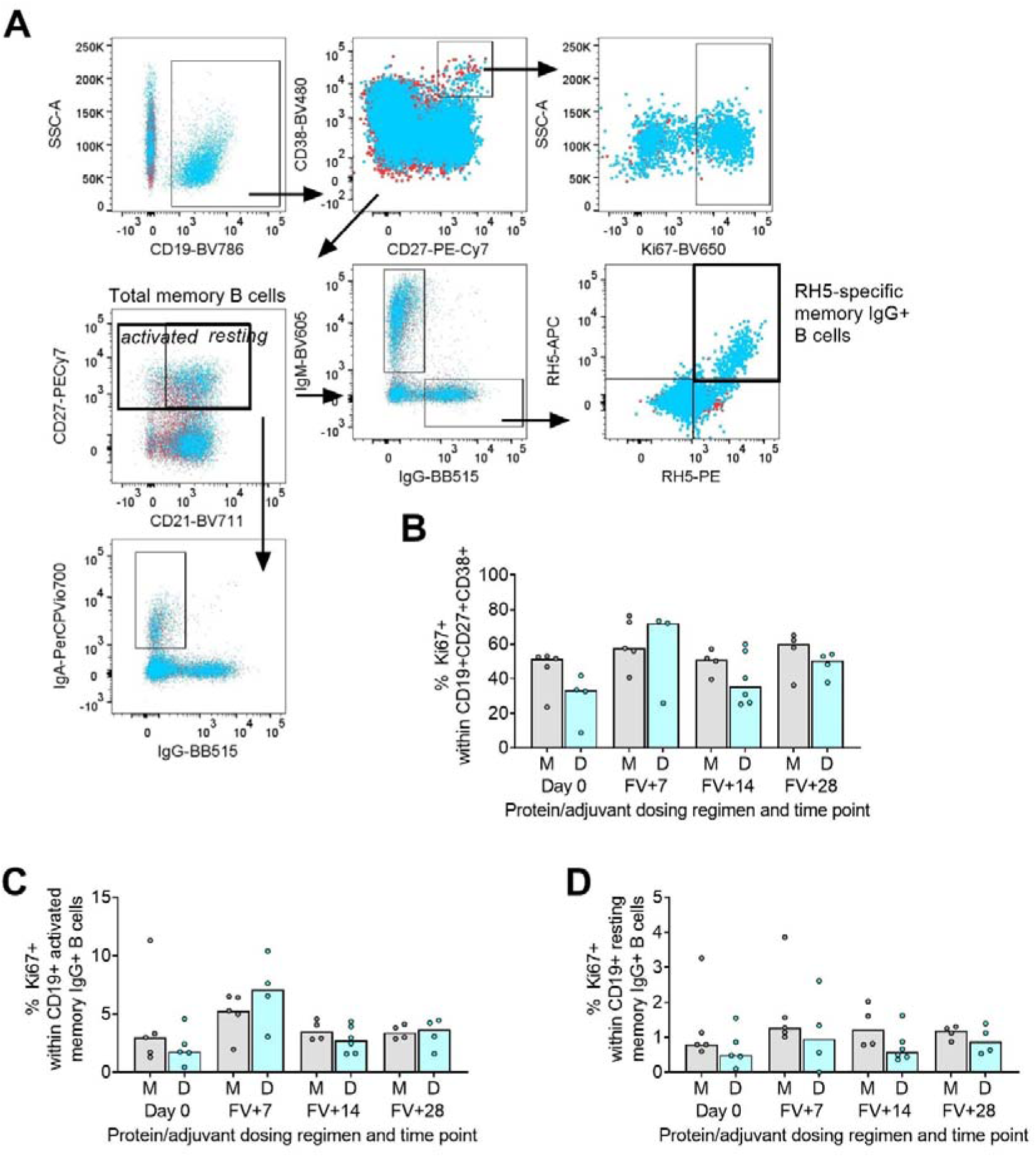
RH5 B cell gating strategy and expression of proliferation marker Ki67. PBMC from pre-vaccination (Day 0) and post-final vaccination (FV) time points were analysed for B cell responses by flow cytometry. (**A**) Gating strategy shows identification of CD19+ B cells within live single (B cell-enriched) lymphocytes, and definition of CD38+CD27+ plasma cells within this population. Total memory cells are defined as CD27+ non-plasma cells (using a NOT gate to exclude plasma cells; indicated with thick black box); activated and resting memory B cells are more specifically categorised as CD21-CD27+ or CD21+CD27+, respectively. IgG+, IgM+ or IgA+ populations are subsequently gated within total memory cells. Proliferating (Ki67+) or vaccine-specific (those co-staining with RH5-PE and RH5-APC probes as indicated by thick black box) cells are defined within the plasma cell or isotype-specific memory B cell populations. Example shows Ki67 expression of plasma cells and RH5-specific gating on memory IgG+ B cells. A FV+14 sample (blue) is overlaid on a matched Day 0 sample (red) for all plots. Frequencies of Ki67+ cells shown within plasma cells (**B**), activated IgG+ memory B cells (**C**), and resting IgG+ memory B cells (**D**). M = RH5.1/adjuvant monthly dosing; D = RH5.1/adjuvant delayed booster dosing. Post-vaccination comparisons were performed between dosing regimens with Mann-Whitney U tests. Sample sizes for all assays were based on sample availability; each circle represents a single sample. (**B-D**) VV/PA: Day 0 = 5/4-5, FV+7 = 5/3-4, FV+14 = 4/6, FV+28 = 4/4. Bars represent medians.

**Supplemental Figure 4.**
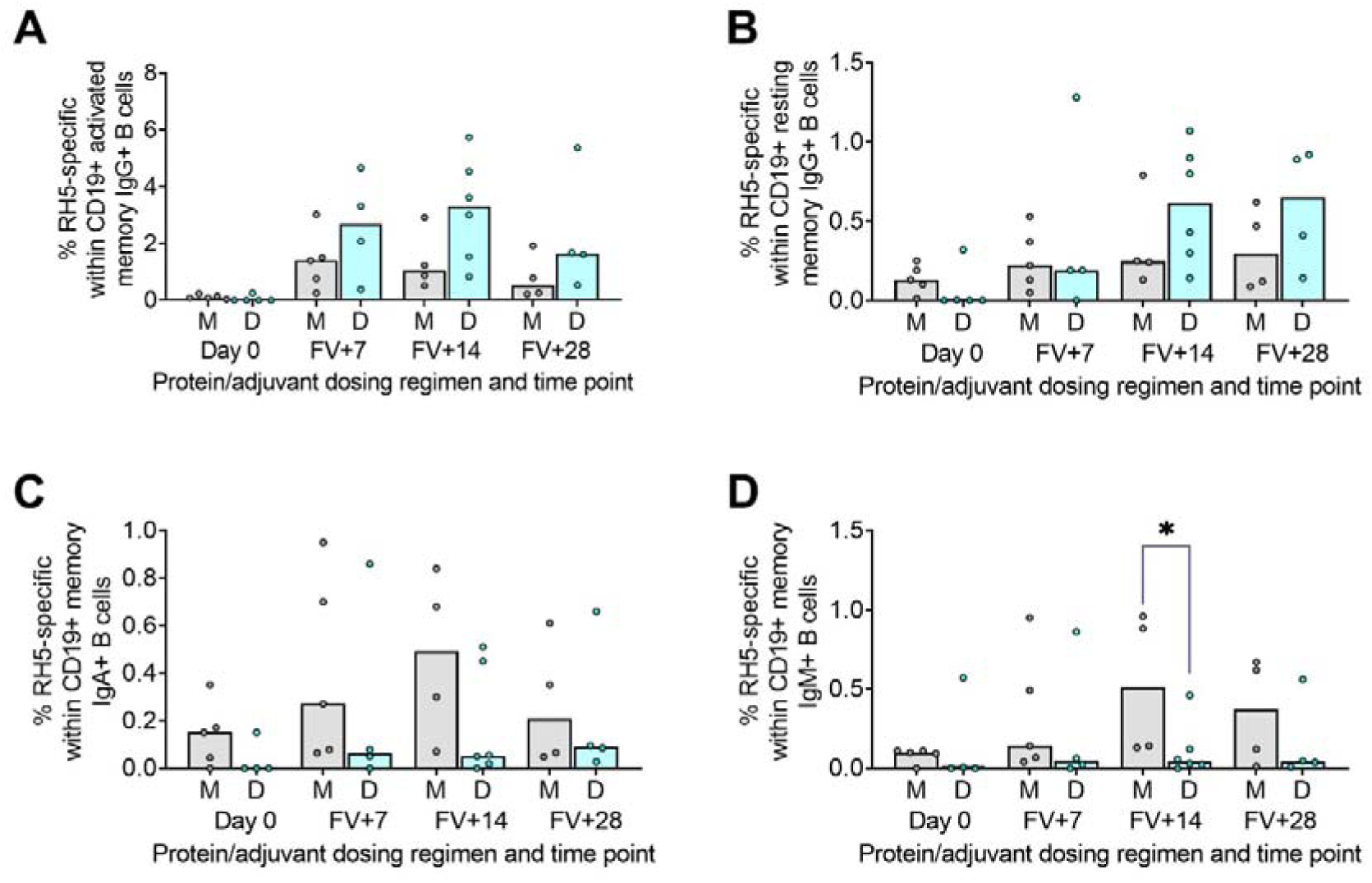
Extended RH5-specific memory B cell responses. PBMC from pre-vaccination (Day 0) and post-final vaccination (FV) time points were analysed for B cell responses by flow cytometry; gating strategies are as described in Methods and **Supplemental Figure 3**. Frequencies of RH5-specific B cells – identified by probe staining – were compared within activated memory IgG+ B cells (**A**), resting memory IgG+ B cells (**B**), total memory IgA+ B cells (**C**), and total memory IgM+ B cells (**D**) between dosing regimens. M = RH5.1/adjuvant monthly dosing; D = RH5.1/adjuvant delayed booster dosing. Post-vaccination comparisons were performed between dosing regimens with Mann-Whitney U tests. Sample sizes for all assays were based on sample availability; each circle represents a single sample. (**B-D**) VV/PA: Day 0 = 5/4-5, FV+7 = 5/4 FV+14 = 4/6, FV+28 = 4/4. Bars represent medians. *\* p* < 0.05.

**Supplemental Figure 5.**
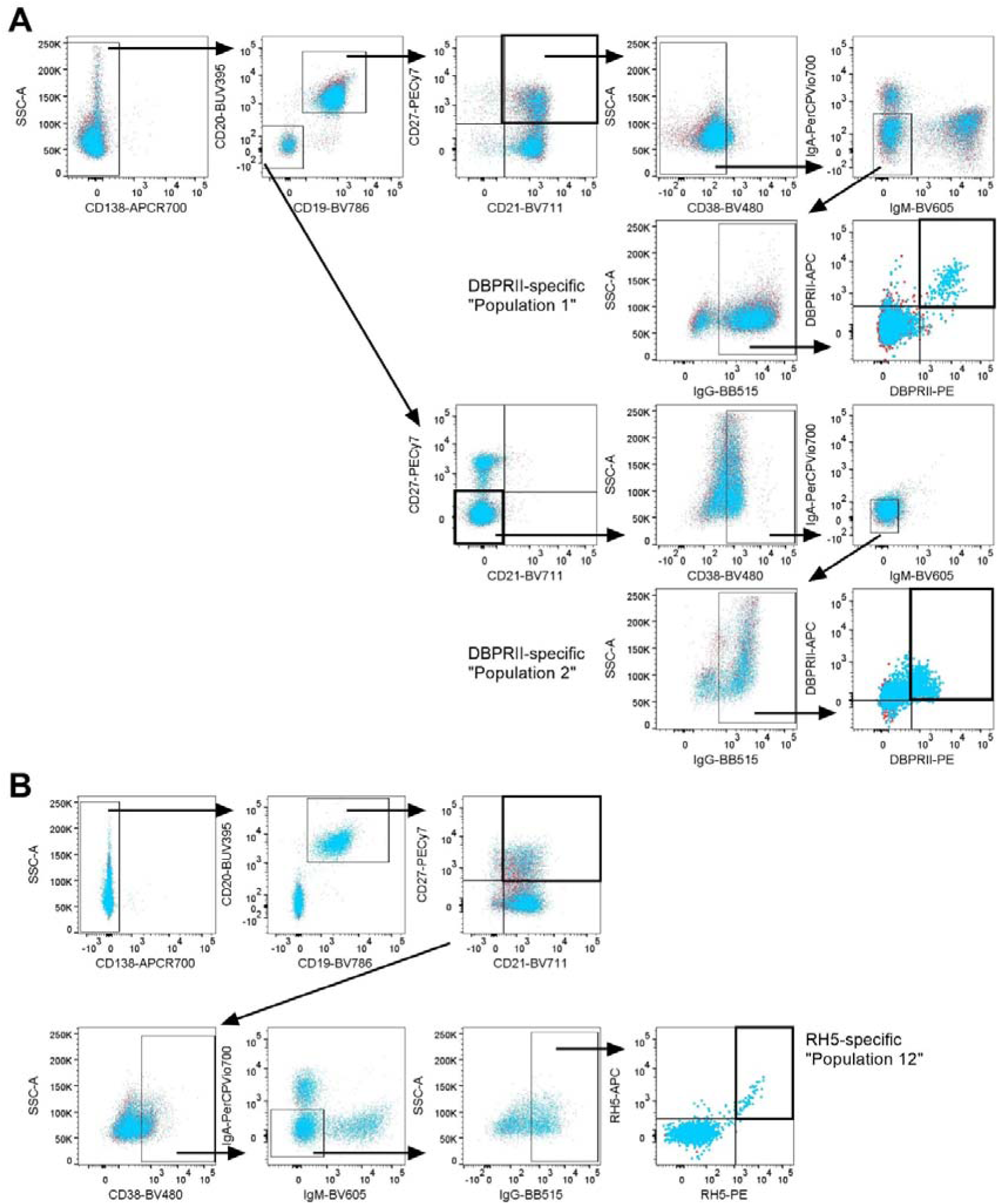
Gating strategies for key agnostically-defined B cell populations via CITRUS. CITRUS was run on single live (B cell-enriched) lymphocyte flow cytometry fcs files to agnostically define the main B cell populations within either DBPRII or RH5 trial samples. Median marker expression within each cluster was used to define gating strategies for B cell populations in FlowJo, which were re-analysed for DBPRII-or RH5-specific responses through probe staining. (**A**) Gating strategy shows identification of “Population 1” (CD19+CD20+CD21+CD27+CD138-CD38-IgM-IgA-IgG+) and “Population 2” (CD19-CD20-CD21-CD27-CD138-CD38+IgM-IgA-IgG+;) within the DBPRII trial and DBPRII-specific cells within these populations. (**B**) Gating strategy shows identification of “Population 12” (CD19+CD20+CD21+CD27+ CD138-CD38+IgA-IgM-IgG+) within the RH5 trial. A FV+14 sample (blue) is overlaid on a matched Day 0 sample (red) for all plots. See **Table 2** for a full list of populations defined via CITRUS.

**Supplemental Figure 6.**
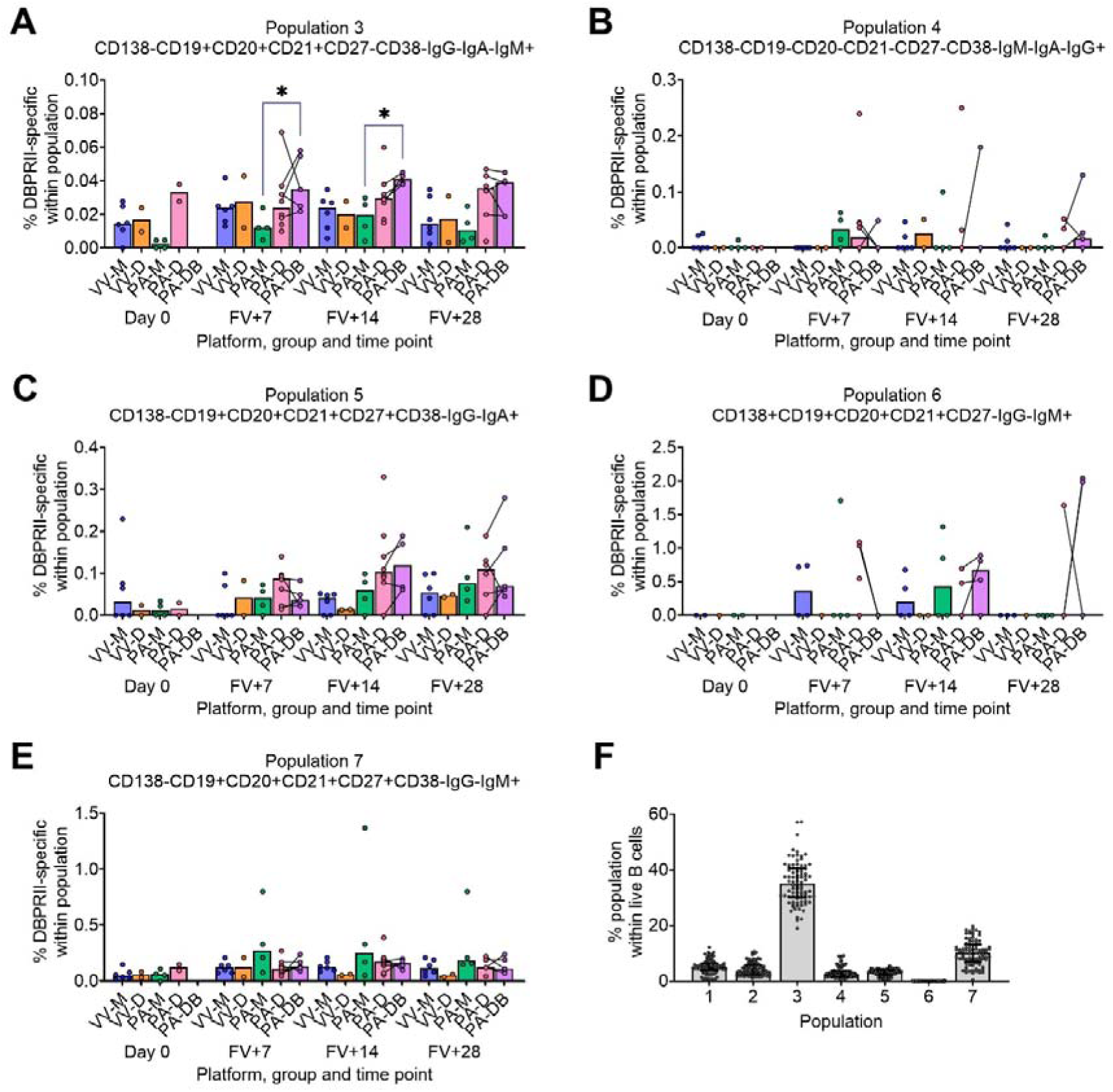
DBPRII-specific responses within agnostically-defined B cell populations. CITRUS was run on single live (B cell-enriched) lymphocyte flow cytometry fcs files to agnostically define the main B cell populations within DBPRII trial samples. Median marker expression within each cluster was used to define gating strategies for B cell populations in FlowJo, which were re-analysed for DBPRII-specific responses through probe staining (**A-E**). Population definitions are annotated on individual figures. (**F**) Frequencies of each population within single live B cells (enriched from lymphocytes; see also **Table 2**) of all samples. VV-M = ChAd63-MVA viral vector monthly dosing; VV-D ChAd63-MVA delayed booster dosing; PA-M = PvDBPII protein/adjuvant monthly dosing; PA-D = PvDBPII protein/adjuvant delayed booster dosing; PA-DB = PvDBPII protein/adjuvant delayed booster dosing with extra booster. FV = final vaccination. Post-vaccination comparisons were performed between PvDBPII protein/adjuvant dosing regimens by Kruskal Wallis test with Dunn’s correction for multiple comparisons (**A-E**). Sample sizes for all assays were based on sample availability; each circle represents a single sample. (**A-C, E**) VV-M/VV-D/PA-M/PA-D/PA-DB: Day 0 = 6/2/4/2/na, FV+7 = 6/2/4/8/5, FV+14 = 6/2/4/8/4, FV+28 = 6/2/4/6/5. PA-D vaccinees returning in the PA-DB group are connected by lines. (**D**) VV-M/VV-D/PA-M/PA-D/PA-DB: Day 0 = 2/1/2/0/na, FV+7 = 4/1/4/8/4, FV+14 = 4/2/4/8/4, FV+28 = 3/1/4/5/5. (**F**) *n* = 86 for all populations. Bars represent medians. * *p* < 0.05.

**Supplemental Figure 7.**
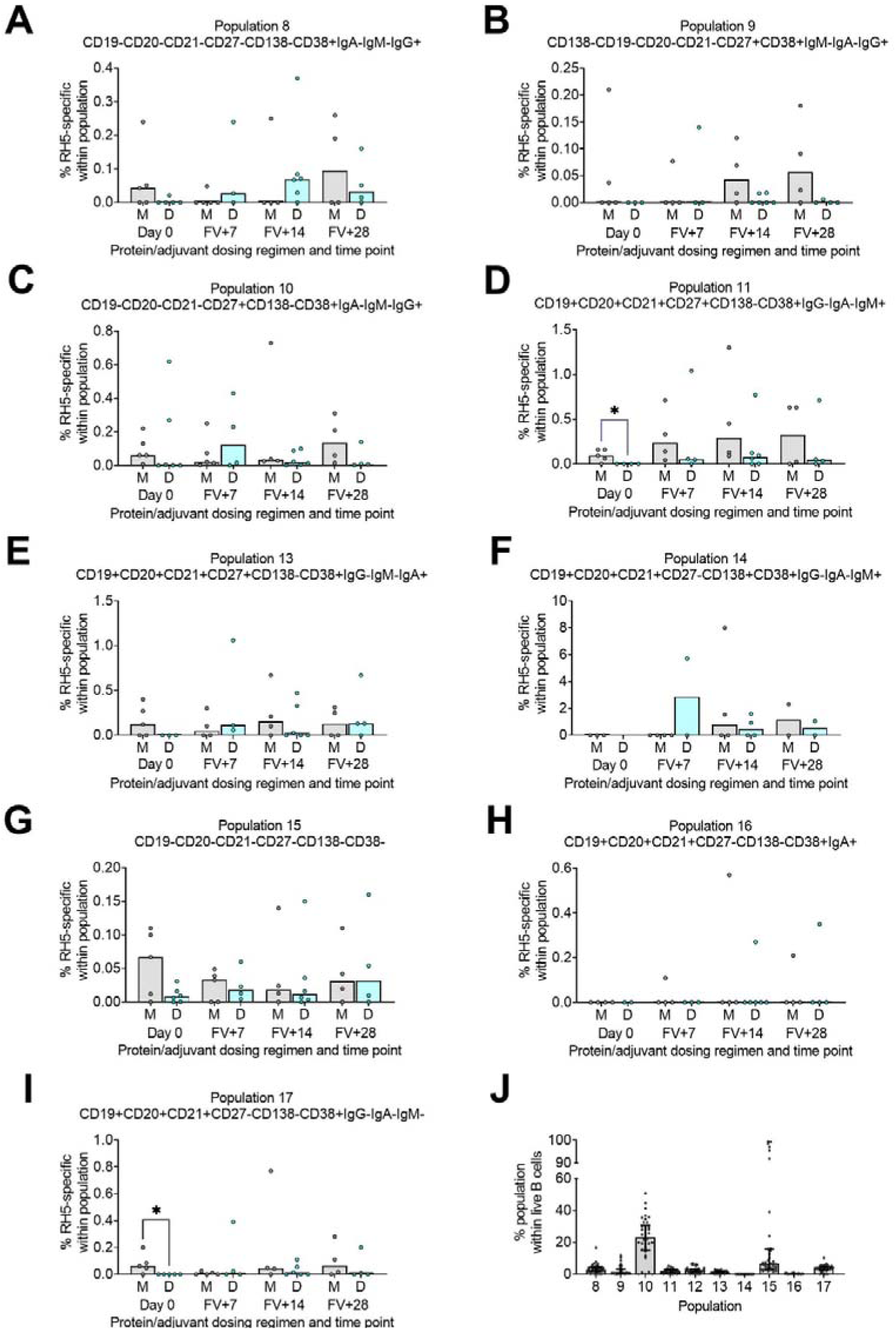
RH5-specific responses within agnostically-defined B cell populations. CITRUS was run on single live (B cell-enriched) lymphocyte flow cytometry fcs files to agnostically define the main B cell populations within RH5 trial samples. Median marker expression within each cluster was used to define gating strategies for B cell populations in FlowJo, which were re-analysed for RH5-specific responses through probe staining (**A-I**). Population definitions are annotated on individual figures. (**J**) Frequencies of each population within single live B cells (enriched from lymphocytes; see also **Table 2**) of all samples. M = RH5.1/adjuvant monthly dosing; D = RH5.1/adjuvant delayed booster dosing. FV = final vaccination. Post-vaccination comparisons were performed between dosing regimens with Mann-Whitney U tests (**A-I**). Sample sizes for all assays were based on sample availability; each circle represents a single sample. (**A-G, I**) M/D Day 0 = 4-5/2-6, FV+7 = 4-5/3-4, FV+14 = 4/6, FV+28 = 4/4. (**H**) M/D Day 0 = 3/0, FV+7 = 4/2, FV+14 = 4/4, FV+28 = 2/2. (**J**) *n* = 38 for all populations. Bars represent medians. * *p* < 0.05.

**Supplemental Figure 8.**
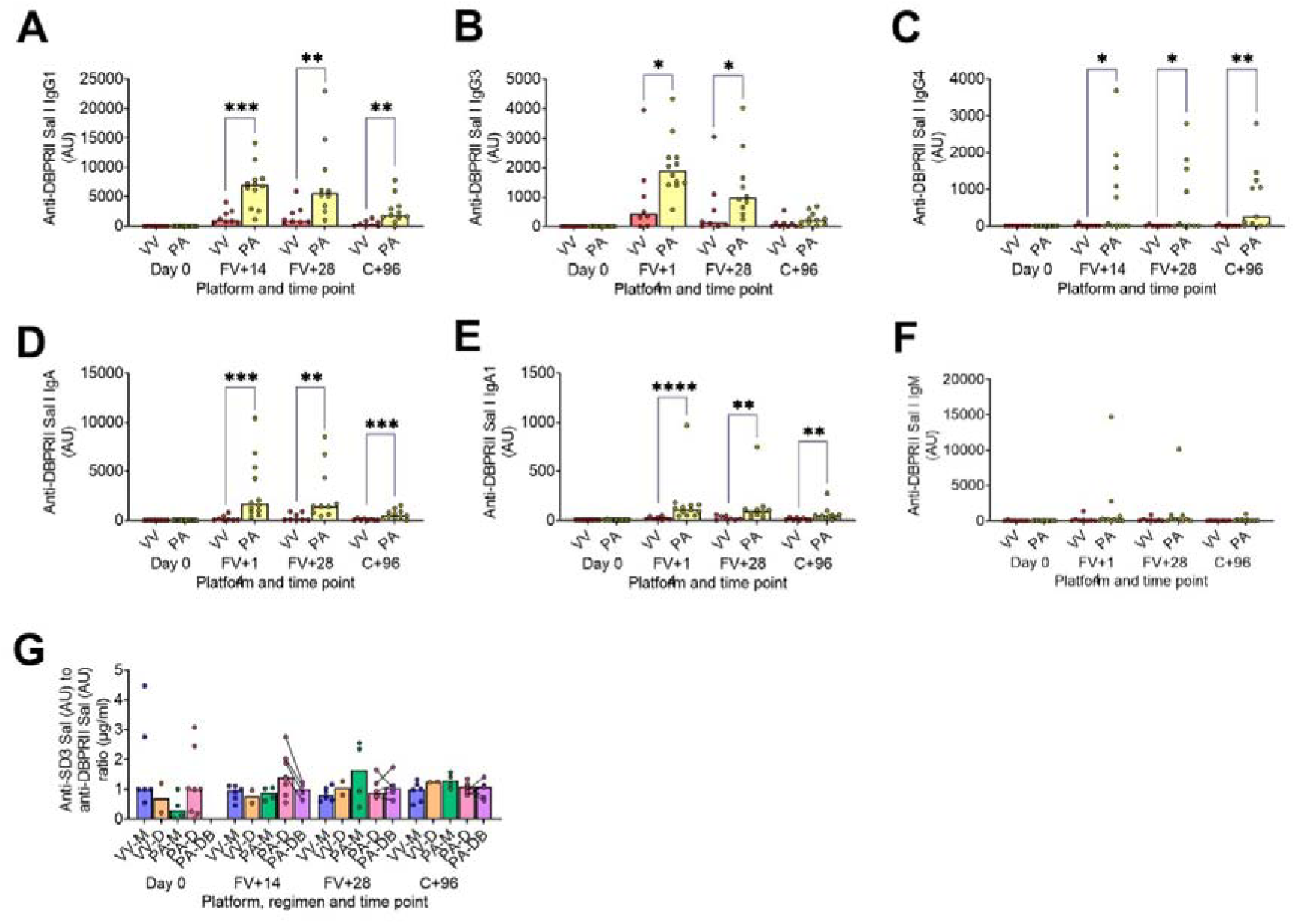
DBPRII-specific peak antibody responses and serum maintenance. Standardised ELISAs were developed to report anti-DBPRII specific or anti-subdomain 3 (sd3) antibody responses in pre-vaccination (Day 0) and post-final vaccination (FV) serum samples. Responses were compared between vaccine platforms for IgG1 (**A**), IgG3 (**B**), IgG4 (**C**), IgA (**D**), IgA1 (**E**), and IgM (**F**). IgG2 and IgA2 responses were below the limit of detection (not shown). The ratio of anti-sd3 to anti-DBPRII was also calculated for total IgG (**G**). VV = ChAd63-MVA viral vectors; PA = PvDBPII protein/adjuvant [PA-M and PA-D]; VV-M = ChAd63-MVA viral vector monthly dosing; VV-D ChAd63-MVA delayed booster dosing; PA-M = protein/adjuvant monthly dosing; PA-D = protein/adjuvant delayed booster dosing; PA-DB = protein/adjuvant delayed booster dosing with extra booster. C+96 = 96 days after controlled human malaria infection (approximately 16-weeks after FV). Post-vaccination comparisons were performed between platforms (**A-F**) with Mann-Whitney U tests, or between protein/adjuvant dosing regimens by Kruskal Wallis test with Dunn’s correction for multiple comparisons (**G**). Sample sizes for all assays were based on sample availability; each circle represents a single sample. (**A-F**) VV/PA: Day 0 = 8/12, FV+14 = 8/12, FV+28 = 8/10. (**G**) VV-M/VV-D/PA-M/PA-D/PA-DB: Day 0 =6/2/4/8/na, FV+14 = 6/2/4/8/4, FV+28 = 6/2/4/6/5, C+96 = 6/2/4/7/5. PA-D vaccinees returning in the PA-DB group are connected by lines. Bars represent medians. * *p* < 0.05, ** *p* < 0.01, *** *p* < 0.001, **** *p* < 0.0001.

**Supplemental Figure 9.**
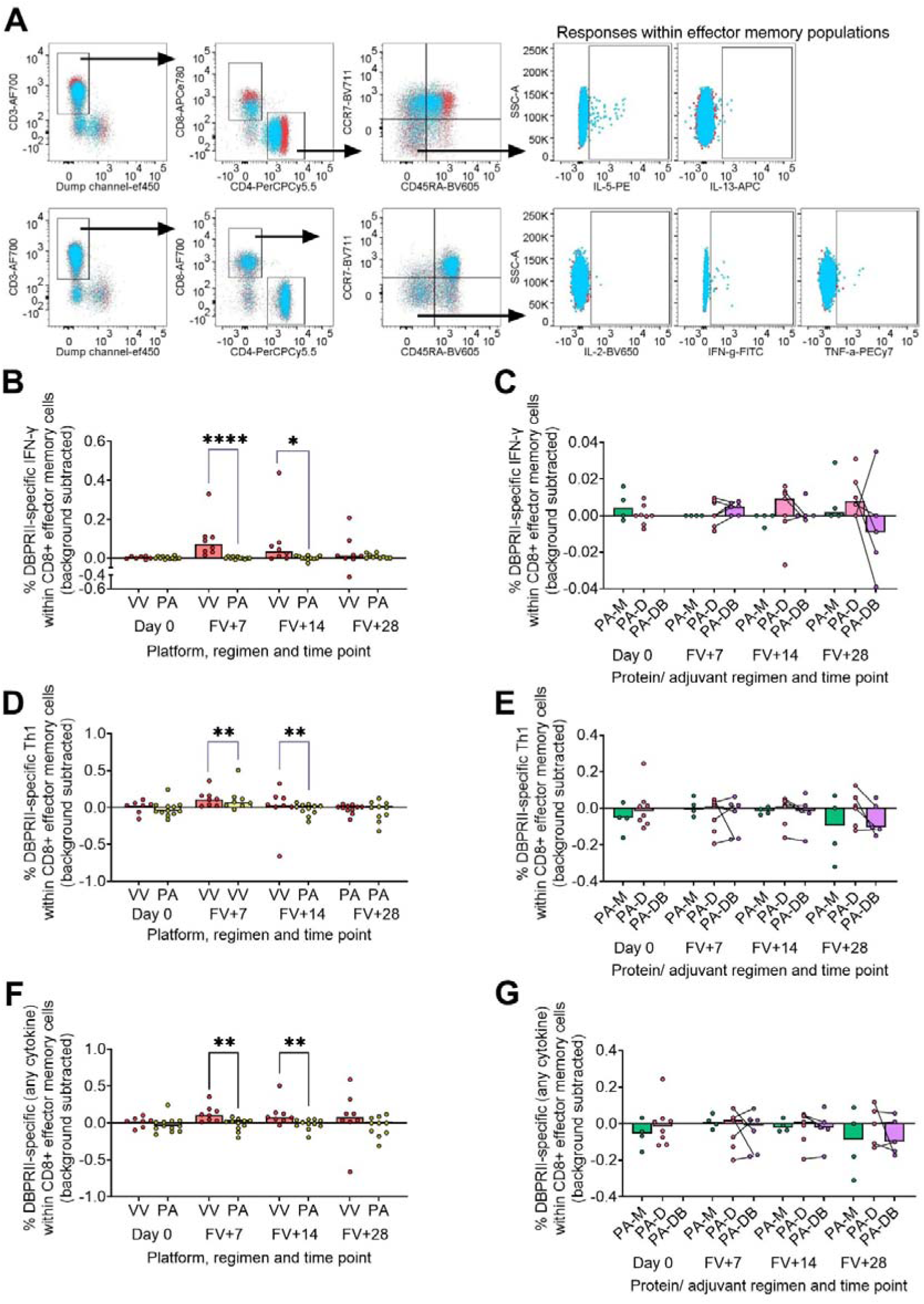
DBPRII-specific T cell gating strategy and CD8+ effector memory responses. PBMC from pre-vaccination (Day 0) and post-final vaccination (FV) time points were analysed for T cell responses by intracellular cytokine staining. (**A**) Gating strategy shows identification of live T cells within single lymphocytes [dump channel includes viability stain, anti-CD14, and anti-CD19; see Methods), and definition of CD4+ and CD8+ T cells within this population. Effector memory CD4+ or CD8+ T cells are identified as CD45RA-CCR7-. Finally, DBPRII-specific T cell responses are defined through detection of intracellular Th2 cytokines (top row; IL-5, IL-13;) or Th1 cytokines (bottom row; IL-2, IFN-γ, TNF-α) following DBPRII peptide pool stimulation. A FV+7 sample (blue) is overlaid on a matched Day 0 sample (red) for all plots (CD4+ effector memory from protein/adjuvant vaccinee used as top row Th2 example; CD8+ effector memory from viral vector vaccinee used as bottom row Th1 example). DBPRII-specific effector memory CD4+ or CD8+ T cells are reported as frequencies producing cytokines in response to peptide stimulation after background subtraction of cytokine-positive cells in matched samples cultured with media alone. Using an ‘OR’ gate, responses were reported for all cytokines (cells producing any of IL-5, IL-13, IL-2, IFN-γ, or TNF-α), Th1 cytokines (IL-2, IFN-γ or TNF-α only), or Th2 cytokine (IL-5 or IL-13 only; see **Figure 4**). CD8+ effector memory T cells IFN-γ (**B-C**), Th1 (**D-E**) or any cytokine (**F-G**) responses were compared between vaccine platforms (**B, D, F**) or protein/adjuvant dosing regimens (**C, E, G**). VV = ChAd63-MVA viral vectors; PA = PvDBPII protein/adjuvant [PA-M and PA-D]; PA-M = PvDBPII protein/adjuvant monthly dosing; PA-D = PvDBPII protein/adjuvant delayed booster dosing; PA-DB = PvDBPII protein/adjuvant delayed booster dosing with extra booster. Post-vaccination comparisons were performed between DBPRII platforms by Mann Whitney U test (**B, D, F**), or protein/adjuvant dosing regimens by Kruskal Wallis test with Dunn’s correction for multiple comparisons (**C, E, G**). Sample sizes for all assays were based on sample availability; each circle represents a single sample. (**B, D, F**) VV/PA: Day 0 = 7/12, FV+7 = 8/11, FV+14 = 8/11, FV+28 = 8/10. (**C, E, G**) PA-M/PA-D/PA-DB: Day 0 = 4/8/na, FV+7 = 4/7/6, FV+14 = 4/7/5, FV+28 = 4/6/5. PA-D vaccinees returning in the PA-DB group are connected by lines. Bars represent medians. * *p* < 0.05, ** *p* < 0.01, **** *p* < 0.0001.

